# Using models to identify the causes of pre-symptomatic transmission from human infection data

**DOI:** 10.1101/2024.05.16.24307410

**Authors:** Kayla Zhang, Damie Pak, Megan A. Greischar

**Affiliations:** Department of Ecology and Evolutionary Biology, Cornell University, 215 Tower Rd, Ithaca, NY 14853, United States

**Keywords:** controlled human infection trial, viral infection, transmission-duration tradeoff, within-host model, norovirus, SARS-CoV-2

## Abstract

When infections can be transmitted from hosts showing no symptoms, containing outbreaks requires distinct strategies like active surveillance. Yet it is rarely clear beforehand when such interventions are needed, especially for emerging pathogens. To investigate the within-host dynamics that enable pre-symptomatic transmission, we survey controlled human infection (CHI) trials with viral pathogens that follow symptoms and viral shedding after inoculation with a known dose. We find that many studies report average timing of symptom onset and shedding, but few report those data for individual participants. We fit a simple model to individual shedding time series from two CHI studies (using norovirus and SARS-CoV-2, respectively) to infer replication rates and the timing of peak shedding relative to symptom onset. We find that faster viral replication significantly hastens peak shedding with minimal impact on symptom onset and no evidence for a tradeoff between the rate and duration of transmission during the pre-symptomatic phase. We then develop and compare within-host models of pathogen replication, immune clearance, and symptom onset to identify plausible assumptions about the causes of pre-symptomatic transmission. We recover the empirical pattern that peak shedding can precede symptom onset when we assume that symptoms are triggered by immune responses rather than pathogen abundance. By incorporating resource limitation via a carrying capacity, we can recover the pattern that faster viral replication prolongs pre-symptomatic transmission. Thus, individual-level data from CHI trials—paired with models—can illuminate the within-host dynamics underpinning pre-symptomatic transmission, guiding efforts to improve control strategies.

**Significance statement:** The COVID-19 pandemic was exacerbated by the potential for transmission before symptoms. Yet the causes of pre-symptomatic transmission—in some hosts but not others—remain unclear, hindering efforts to predict disease spread and tailor control efforts for novel pathogens. To identify patterns across viral taxa, we surveyed controlled human infection (CHI) trials, which rarely reported data on the onset of shedding and symptoms for individuals. Individual time series of shedding and symptoms were available from two CHI trials using norovirus and SARS-CoV-2. We fit models to those data to show that faster viral replication hastens shedding but not symptom onset and then used more detailed models to identify plausible assumptions about the within-host causes of underlying pre-symptomatic transmission.

## 1 Introduction

Controlling outbreaks is more difficult when infections can be transmitted from hosts showing no symptoms, either prior to symptom onset (pre-symptomatic transmission) or without symptoms (asymptomatic transmission) [3]. The COVID-19 pandemic exemplifies this challenge, since up to an estimated 69 percent of transmission was from pre-symptomatic hosts [4]. In contrast, SARS-CoV demonstrated far less pre-symptomatic transmission and was contained [3]. The variation in pre-symptomatic transmission, even among related pathogens, makes it difficult to anticipate how best to control emerging pathogens. Hence recent theory has focused on when and why pathogens evolve pre-symptomatic transmission [5–7]. Yet anticipating pre-symptomatic transmission remains difficult due to lack of clarity about the within-host dynamics that govern the relative timing of host infectiousness and symptoms.

Pathogens’ potential rate of transmission is classically assumed to scale with their rate of replication within the host [8]. That assumption finds support in positive correlations between infectiousness and pathogen load observed in a range of systems (reviewed in [9]), including HIV [10] and dengue [11]. Since symptoms are ultimately a consequence of pathogen replication, symptom onset might also be expected to occur earlier with faster replication rates. Classic theory predicts that traits that enhance transmission—like faster replication and greater within-host abundance—should also come at the cost of truncating infectiousness by killing the host, constraining pathogen evolution to intermediate, rather than maximal, rates of host exploitation [8]. Symptoms could serve a similar function if symptomatic hosts are less likely to contact other hosts. However, the relationship between pathogen load and symptoms is less clear, except in extreme cases such as HIV, where individuals with higher viral loads progress to AIDS—and its associated symptoms—more quickly [10]. Symptoms could be directly correlated to pathogen load, for example through toxins secreted by pathogens [12]. In these cases, faster within-host replication and subsequently greater pathogen loads should hasten disease progression, including the onset of symptoms, as it does in HIV [10, 13]. Alternatively, symptoms could be triggered by the immune response, e.g., fever [14]. Faster pathogen replication here could hasten both symptoms and clearance through upregulation of immune defenses, the latter of which has been shown in dengue [15]. Without a clear understanding of the links between symptom onset and transmission, it is impossible to anticipate constraints on pathogen evolution.

Distinct from the role of replication rates, the relative timing of symptom onset and transmission could also be impacted by initial dose. A higher inoculum dose should reduce the time required for pathogens to replicate up to transmissible densities within the host, but the consequences for symptom onset are less clear. A higher dose could reduce pre-symptomatic transmission by triggering an immune response more quickly or increasing the initial amount of pathogen secretions within the host, leading to the rapid onset of immune- or pathogen-mediated symptoms. In contrast, lower inoculum doses may prevent activation of the host immune response across bacterial pathogens [16], delaying any immune-mediated symptoms. Recent analysis of norovirus data suggests that larger doses may be associated with earlier shedding onset and peak, as well as hastened symptom onset [1]. Thus, the impacts of dose on the relative timing of symptom onset and shedding warrant more attention.

To investigate the causes of pre-symptomatic transmission, we surveyed controlled human infection (CHI) trials of viral pathogens for data on the timing of onset for symptoms and transmission (that is, viral shedding). In CHI trials, volunteers are inoculated with a known dose of a pathogenic organism deemed sufficiently low-risk [17], providing precise data on inoculum dose, and the timing of shedding and symptoms (e.g., [18]) that cannot be obtained from observation of natural infections. We found that few CHI studies reported the timing of symptoms and shedding for individual infections and subsequently focused on two studies with detailed time series data for individuals, a re-analysis of a CHI trial with norovirus [1, 19], and a recent SARS-CoV-2 trial [2]. To identify how pre-symptomatic transmission varies with inoculum dose and viral replication rates, we fit a statistical model [20] to viral shedding data to estimate replication rates and the time of peak shedding for individual infections. We classified infections as showing pre-symptomatic transmission when the estimated peak in pathogen shedding occurred prior to symptom onset for convenience of later comparison to within-host models. We found no strong evidence for dose-dependence in norovirus infections, nor evidence in either virus for a tradeoff between the rate and duration of pre-symptomatic transmission that is commonly assumed in theory [5–7, 21]. Faster replication rates hastened peak shedding without changing symptom timing, prolonging pre-symptomatic transmission. By modifying a simple model of pathogen replication and removal by immune effectors [22], we identified assumptions about viral replication and symptom onset consistent with empirical patterns. We found that peak shedding could only precede symptom onset when we assumed symptoms were triggered by increasing abundance of immune effectors, rather than pathogen load. We recovered a positive relationship between viral replication rate and pre-symptomatic transmission when we incorporated a pathogen carrying capacity, with our analysis suggesting that immune inhibition of pathogen replication is insufficient to recover empirical patterns. Thus, individual data on pathogen shedding and symptom onset enable selection of plausible models for pre-symptomatic transmission.

## 2 Results

We first review of CHI trials to survey what data are reported regarding the timing of symptoms and transmission. We then use a statistical model [20] to estimate the timing of peak viral shedding as well as pathogen replication rates to determine trends in pre-symptomatic transmission among patients in a norovirus [1] and SARS-CoV-2 study [2], two of seven studies that included individual data on onset of both viral shedding and symptoms, and two of four that made available individual time series of shedding. Lastly, we modify a dynamical model [22] to identify biological assumptions that give rise to empirical patterns.

### 2.1 Most CHI trials do not report individual timing of shedding & symptom onset

We find 41 CHI studies reporting the onset time for both symptoms and shedding, summarized in Fig. 1, including rhinovirus [25, 27–37] (Table S1), influenza A [23–25, 38–43] (Table S2), SARS-CoV-2 [2, 44] (Table S3), respiratory syncytial virus (RSV) [26, 45–58] (Table S4), and norovirus [1, 18, 59, 60] (Table S5). All influenza A studies (save one) and both SARS-CoV-2 studies exhibit a pattern of viral shedding preceding symptom onset, when averaged across participants. Rhinovirus, RSV and norovirus exhibit substantial variation in the average relative timing of symptoms versus shedding (Fig. 1). Only 7 studies report the timing of symptom and shedding onset for individuals [1, 2, 18, 23–26]. Four of those seven studies also report individual-level viral shedding time series data [1, 2, 25, 26], revealing considerable variation across individuals beyond what can be explained by differences in dose [1]. While some viruses show repeatable patterns in average timing of shedding and symptoms—leaving open the possibility of a phylogenetic signal—there is substantial variation across and within-studies. We next use individual shedding data from norovirus [1] and SARS-CoV-2 [2] to determine the extent to which this variation can be explained by within-host replication rates.

**Figure 1:**
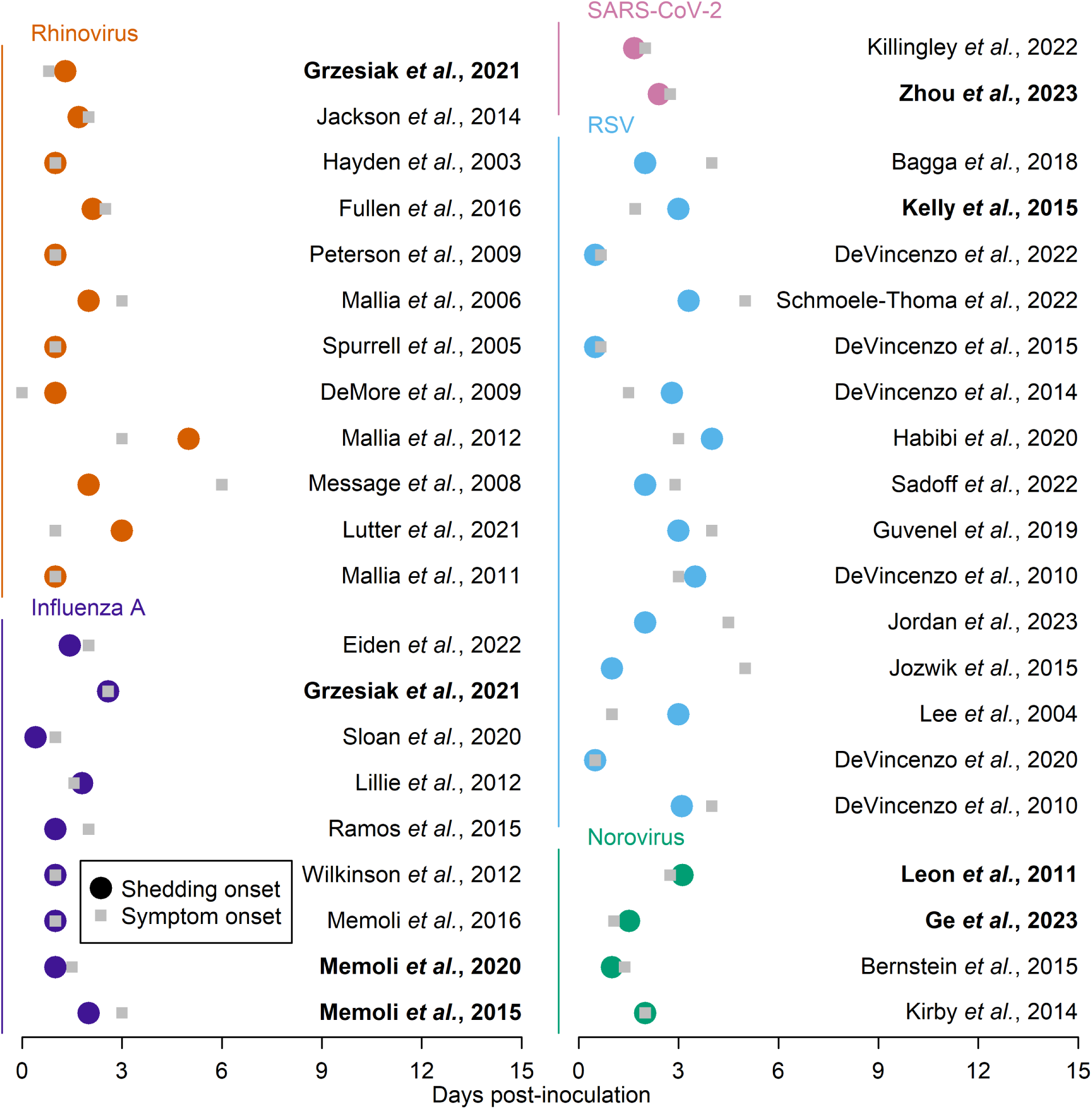
Most CHI studies do not report individual-level data on both pathogen shedding onset and symptom onset. Circles indicate shedding onset, while squares indicate symptom onset. The 7 studies that report individual-level data for both shedding and symptom onset are bolded [1, 2, 18, 23–26]. Note that [1] is a re-analysis of the data from [19]. Sixty-five CHI studies were originally identified as having data on either shedding or symptoms, but only 41 (shown here) included data on both.

### 2.2 Faster viral replication prolongs pre-symptomatic transmis-sion

We use the viral shedding data reported by [1] (a re-analysis of the data from [19]) and [2] to reconstruct the within-host trajectory of viral load by fitting the rates of exponential growth and decline, and the timing and height of peak viral load for each individual in each study. For simplicity, we assume that shedding time series reflect within-host dynamics with no time delay and, by extension, no variation in the time delay between within-host dynamics and shedding across hosts. We later discuss how a lag between viral load and shedding could impact our results and conclusions. To estimate the trajectory of viral load, we fit a statistical model [20] to individual viral shedding data (fitted parameters in Table S7). The log_10_ viral load, *V* (*t*), is defined as

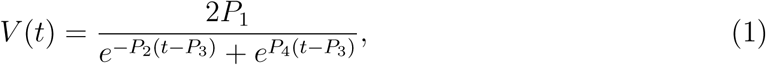

where *P*_1_ represents the log_10_ of peak viral load; *P*_2_ and *P*_4_ refer to the exponential growth and decay rates, respectively, of viral load, and *P*_3_ refers to the time of peak viral load.

We initially define pre-symptomatic transmission as occurring when peak viral shedding preceded symptom onset, on the logic that individuals shedding at their maximal rates prior to symptoms were likely to be infectious and to enable comparison to within-host models. When fit to data from [1] and [2], estimated individual trajectories show variation in the relative timing of peak shedding and symptom onset (Fig. 2, data and fitted trajectories for other patients in Figs. S1 and S2). Viral shedding typically increases initially, then decreases, with the estimated time of peak shedding (*P*_3_ in Eq. 1) either preceding or following symptom onset. For example, of the three individual infections shown in Fig. 2, only one (B) exhibits pre-symptomatic transmission.

**Figure 2:**
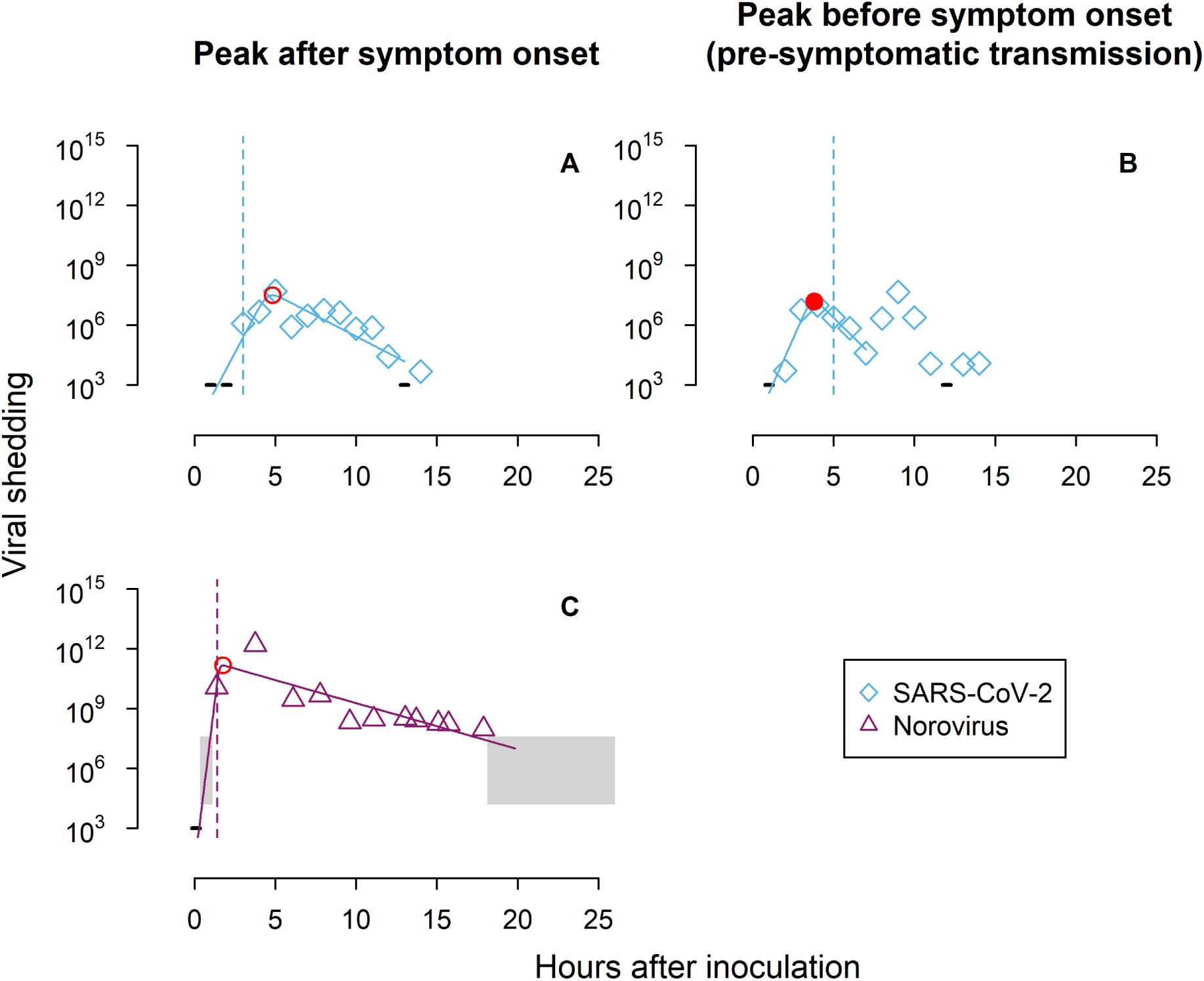
The relative timing of symptoms and shedding varies across individuals. Panels A, B, and C show viral shedding over time for two participants (Throat 16 and Throat 5) infected with SARS-CoV-2 from [2] and one participant (ID13) infected with norovirus from [1]. Open diamonds or triangles indicate observed viral shedding in the hours after inoculation with a solid line to indicate the best fit parameterization of Eq. 1 to the data. The dotted vertical lines denote symptom onset. A red circle shows estimated peak viral shedding; a closed red circle indicates peak shedding preceded symptoms (pre-symptomatic transmission, B only), and open red circles show peak shedding following symptom onset (post-symptomatic transmission, A and C). A grey rectangle indicates the range of shedding values for samples that fall between the detection limits for two different methods (norovirus only), while horizontal black lines indicate samples where no virus was detected.

Across all individuals with a clear peak in shedding and who exhibited symptoms during the study, none of the participants of the norovirus study exhibit pre-symptomatic trans-mission, such that peak shedding preceded symptom onset. While not apparent from the data reported by [1], past CHI data suggest that peak shedding can precede symptom onset in norovirus infections [19] (Fig. S3). In the norovirus data, there is no clear relationship between dose and estimated replication rates or time of peak shedding, but there is a (non-significant) negative correlation between dose and time of symptom onset (Fig. S4), such that increasing doses delay peak shedding relative to symptom onset (Fig. 3A). The SARS-CoV-2 study focused on a single dose, where three infections exhibit a peak in shedding preceding symptoms. We find a significant negative correlation between estimated viral replication rate (*P*_2_ parameter in Equation 1) and the delay between symptom onset and peak viral shedding for norovirus participants (Fig. 3B, p = 0.00060*, β* = *−*1.5*, R*^2^ = 0.56), with a similar correlation in the SARS-CoV-2 data (*p* = 0.24*, β* = *−*2.4*, R*^2^ = 0.15). Thus, faster replication prolongs pre-symptomatic transmission, since higher replication rates lead to significantly earlier peak shedding (*p* = 4.0 *×* 10*^−^*^7^*, R*^2^ = 0.83 for norovirus, *p* = 5.34 *×* 10*^−^*^6^*, R*^2^ = 0.88 for SARS-CoV-2) while having no marked impact on symptom onset (Fig. 3C-D). When we instead define pre-symptomatic transmission less conservatively as shedding onset preceding symptom onset, we find that greater doses again slightly prolong shedding onset relative to symptoms, though the effect is more subtle than if we focus on peak shedding, Fig. S6A). We find a similar pattern such that faster replication hastens shedding onset, though that correlation is weaker than for the peak in shedding (significant for norovirus, *p* = 0.0061*, R*^2^ = 0.40, nonsignificant for SARS-CoV-2, *p* = 0.28*, R*^2^ = 0.12). As a result, we no longer find a significant net effect of replication rates on the timing of shedding versus symptom onset (Fig. S6B-D).

**Figure 3:**
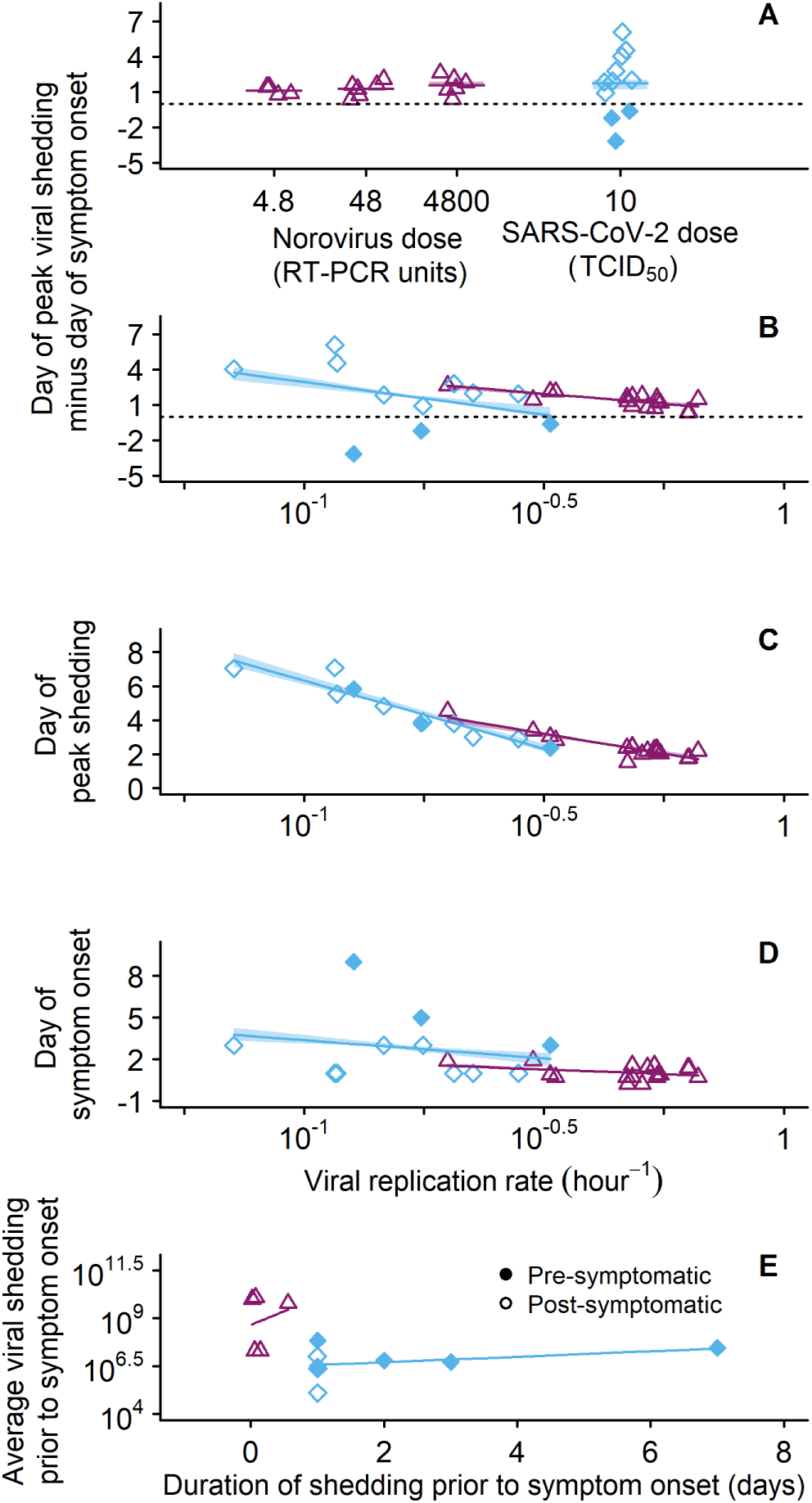
Individual-level data suggest that faster replication prolongs pre-symptomatic transmission. (A) Pre-symptomatic transmission was observed for SARS-CoV-2 (blue diamonds, [2]) but not norovirus (purple triangles, [1]). Horizontal bars indicate the average delay between peak viral shedding and symptom onset for each dose (only one dose was used in the SARS-CoV-2 study), with a horizontal dotted line to indicate zero (simultaneous peak shedding and symptom onset) in panels A and B. (B) Faster replication rates reduce the delay between symptom onset and peak shedding (*P*_2_, Eq. 1, *p* = 0.00060*, R*^2^ = 0.56 for norovirus, *p* = 0.24*, R*^2^ = 0.15 for SARS-CoV-2). (C) Faster replication hastens peak shedding for both viruses (*p* = 4.0 *×* 10*^−^*^7^*, R*^2^ = 0.83 for norovirus, *p* = 5.34 *×* 10*^−^*^6^*, R*^2^ = 0.88 for SARS-CoV-2). (D) There is no significant correlation between the timing of symptom onset and pathogen replication rate (*p* = 0.18*, R*^2^ = 0.12 for norovirus, *p* = 0.55*, R*^2^ = 0.041 for SARS-CoV-2). (E) There is also no clear relationship between average transmission rate (i.e., viral concentration averaged across time points) prior to symptoms and the duration of pre-symptomatic transmission (symptom onset minus shedding onset). Shaded regions represent the 95% confidence intervals obtained by bootstrapping via case resampling of 100 parameter fits per individual infection (omitted in panel E due to small number of points).

We also calculate average shedding prior to symptom onset (viral concentration averaged across sampling times) for each participant as a proxy for transmission rate. Theory often assumes a tradeoff between the duration and rate of transmission prior to symptoms (e.g., [5]), but we find no such negative correlation between the duration of shedding prior to symptoms and average shedding using linear regression (Fig. 3E). When we repeat this test using average total shedding (viral concentration times sample weight averaged across sampling times) for norovirus, we still find no correlation (Fig. S5).

### 2.3 An immune symptom threshold and pathogen carrying capacity can reproduce empirical patterns

We next compared within-host models to identify assumptions that can recapitulate the two main patterns identified from the data: (1) that peak shedding can precede symptom onset (pre-symptomatic transmission); and (2) that faster viral replication rates prolong pre-symptomatic transmission. We extended a simple model of exponential pathogen replication and removal by immune effectors [22] to incorporate symptom onset (“base model”), based either a pathogen load threshold (*s_P_*), or an immune effector threshold (*s_X_*). Assuming a pathogen load threshold, peak viral load can never precede symptom onset. If peak viral load falls below the symptom threshold *s_P_*, then the simulated infection will never cause symptoms (Fig. 4A, left). Alternately, if peak viral load is greater than the symptom threshold *s_P_*, then viral load must cross the threshold before it peaks (Fig. 4A, right). Because peak viral load can never precede symptom onset, pre-symptomatic transmission as seen in the data is not possible, only post-symptomatic and asymptomatic transmission.

**Figure 4:**
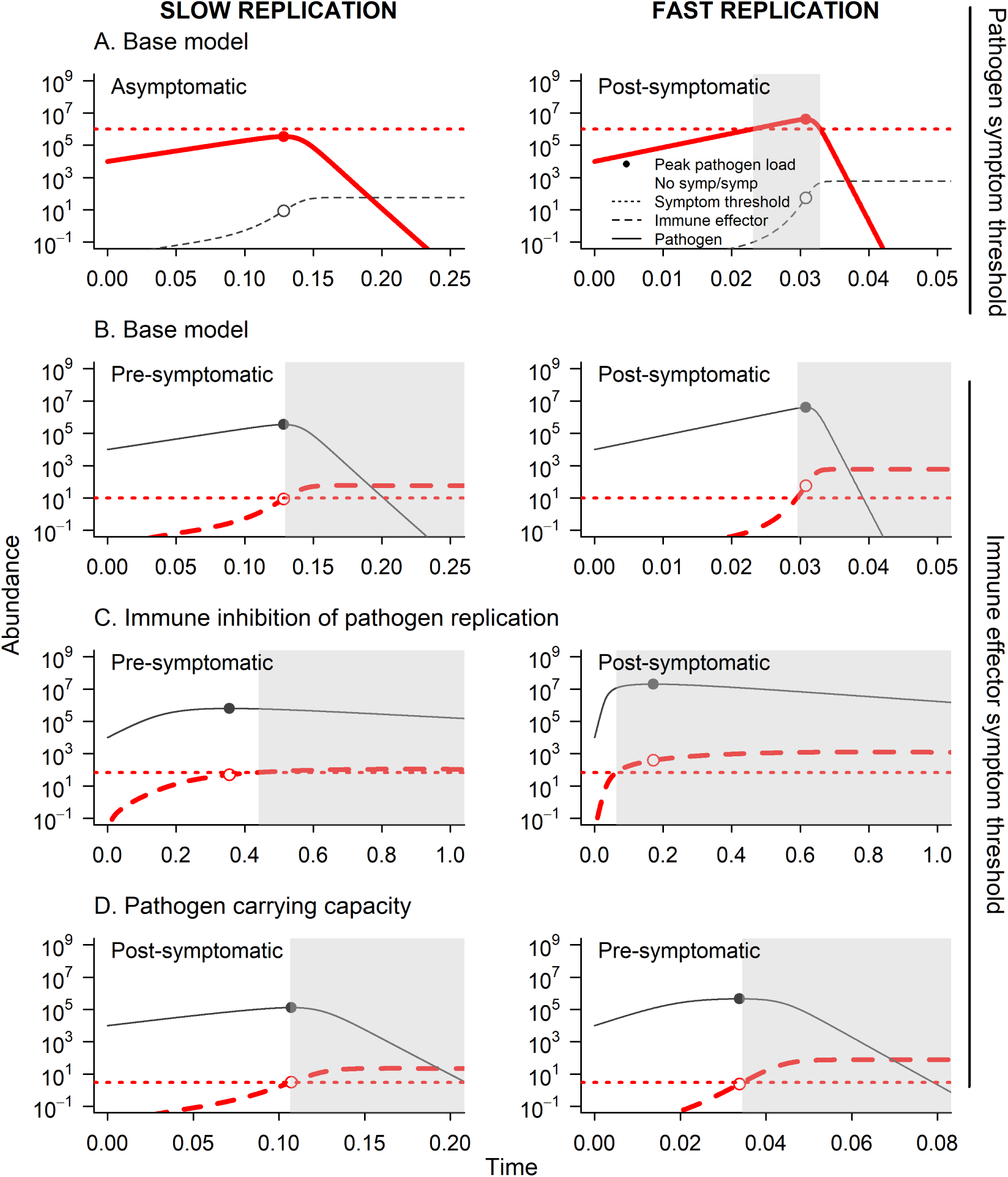
Models reveal plausible (and implausible) drivers of pre-symptomatic transmission. Bolded red lines indicate the quantity that triggers symptoms when its abundance exceeds the threshold (dotted horizontal line). Pathogen abundance is indicates with a solid curve (and closed dot at the peak), while immune effector abundance is shown as a dashed line with an open circle at peak pathogen abundance. (A) With a pathogen abundance symptom threshold (*s_P_* = 10^6^), the base model (Eqns. 2) can only produce asymptomatic or post-symptomatic transmission (when replication is slow or fast, respectively). (B) With an immune effector abundance symptom threshold (*s_X_* = 10), the model can produce pre- and post-symptomatic transmission, with slow and fast replication (respectively). (C) Retaining an immune effector abundance symptom threshold (*s_X_* = 70) and assuming that immunity interferes with pathogen replication rather than direct killing, and that immune upregulation saturates at a certain level (Eqns. 3), pre-symptomatic transmission is more likely with slow replication. (D) Retaining an immune effector abundance symptom threshold (*s_X_* = 3), assuming direct killing of pathogens, and relaxing the assumption of exponential pathogen growth (Eqns. 4), pre-symptomatic transmission is more likely with faster replication. Replication rates were *r* = 30 (left panels) or *r* = 200 (right panels).

When we modified this base model to assume that symptoms occur when immune effector abundance exceeds a threshold, both pre-symptomatic and post-symptomatic transmission were possible (i.e., peak pathogen abundance could precede or follow symptom onset, Fig. 4B). However, our analysis showed that as replication rate *r* increases, the simulated peak in pathogen abundance switched from pre-to post-symptom onset, a pattern we confirmed with simulations (Fig. S7A). We confirmed qualitatively identical behavior for a model with immune inhibition of pathogen replication rather than direct removal of pathogen. In both models, the timing of peak pathogen abundance for a given replication rate (*r*) is a function of immune effector abundance (see Methods for details). Higher replication rates require a larger immune effector abundance to force a peak and subsequent decline in pathogen abundance. Faster replication then makes it more likely that immune effector abundance will cross a symptom threshold (*s_x_*) prior to peak pathogen abundance (Fig. S8A). Thus, the models that assume immunity is the only check on pathogen replication fail to recover the empirical pattern that faster replication extends pre-symptomatic transmission (Fig. 3B).

We then investigated the impact of incorporating a pathogen carrying capacity on pre-symptomatic transmission, implying resource-limitation, so that immune clearance is not the sole check on pathogen replication. We again found that peak pathogen abundance could precede or follow symptom onset (both pre- and post-symptomatic transmission, Fig. 4C). By relaxing the assumption of exponential pathogen growth, we recovered the empirical pattern that as replication rate increases, there is a longer period of time between peak pathogen load and symptom onset (pre-symptomatic transmission, Fig. S7B, *cf.* Fig. 3B, Fig. S8B). In this model, the timing of peak pathogen abundance depends on both immune effector and pathogen abundance (see Methods for details), so that faster replication can hasten peak timing without also speeding up symptom onset.

## 3 Discussion

Pre-symptomatic transmission plays an important role in propagating outbreaks of diseases, including SARS-CoV-2 (e.g., [61]). Better understanding of how within-host dynamics determine the relative timing of transmission and symptoms would improve prevention strategies by enabling predictions about the host and pathogen characteristics likely to promote pre-symptomatic transmission. We showed that achieving that level of understanding could be greatly facilitated by applying within-host models to the invaluable data generated by CHI trials. We found that individual data on shedding and the timing of symptoms are rarely reported from viral CHI trials, but those data can be leveraged to pare down the proximate and evolutionary drivers of pre-symptomatic transmission. By analyzing previously reported data [1, 2], we uncovered that faster within-host replication hastens peak shedding with minimal impact on symptom onset. We found no strong evidence for dose-dependence or for a tradeoff between transmission rate and duration during pre-symptomatic infection. Subsequent analysis revealed that the model most consistent with the data was one in which symptom onset is triggered by immune effector abundance exceeding a threshold and in which pathogen populations are regulated by both resource limitation and immune clearance. Our study provides proof-of-concept for how within-host models can extend the inferences drawn from CHI trials.

We found that pathogen replication rate was positively associated with earlier peak shedding prior to symptom onset. That pattern was similar for norovirus and SARS-CoV-2— two highly divergent viruses–suggesting that such a pattern could play out across pathogen species, with faster-replication enhancing pre-symptomatic transmission. SARS-CoV-2 and H1N1pdm09 (the latter responsible for the 2009 influenza pandemic) display early peak viral loads and appreciable amounts of pre-symptomatic transmission, while SARS-CoV and MERS-CoV exhibit later peak viral loads and little pre-symptomatic transmission [62]. Individual-level data from CHI trials could reveal whether this pattern holds across diverse pathogens and whether there is any phylogenetic signal of pre-symptomatic transmission. If patterns of pre-symptomatic transmission are phylogenetically conserved, it would enhance capacity to predict the spread of emerging pathogens. Distinct from any phylogenetic patterns, predictive ability would also be improved by identifying tradeoffs that constrain the evolution of pre-symptomatic transmission.

In contrast to our finding that faster replication hastens peak shedding, we found consid-erable variability in the timing of shedding onset and no clear relationship with replication rates. We focused on peak shedding due in part to the difficulty of interpreting how early shedding relates to transmission success. For the norovirus study, viral shedding was quantified by PCR [1, 19], which could detect inviable virus, while the SARS-CoV-2 shedding data were obtained by plaque assays [2] that detect viable virus only. Even when shedding data account for viability, two unknowns make it difficult to interpret the significance of early shedding: 1) how dose impacts the probability of infecting another host, including whether there is a minimum required inoculation dose; and 2) investigation of what inoculation doses are typical for different pathogenic organisms. Some of these unknowns have been investigated in other human viral infections, including experimental studies that map viral load to the probability of transmission in HIV and dengue [9]. Estimates of infective dose have likewise been used to quantify implications for virulence [63]. Identifying exposure to viruses is challenging, but progress has been made for SARS-CoV-2, for example, using models of exhalation flow physics to quantify how mask-wearing reduces exposure [64]. Progress in these areas would enhance capacity to understand how pre-peak shedding contributes to infectiousness, including following interventions designed to disrupt transmission.

Our analysis provides no evidence for classic tradeoffs between the rate and duration of transmission. Faster replication promoted pre-symptomatic transmission by hastening peak pathogen shedding, but had no impact on the timing of symptom onset (Fig. 3C-D). Moreover, there was no relationship between transmission rate (i.e., both viral concentration and total shedding averaged over the samples) prior to symptoms and the duration of this pre-symptomatic shedding, which belies the expected tradeoff between transmission and duration for this initial latent phase of infection, commonly assumed in theory [5–7, 21]. While such a transmission-duration tradeoff is highly relevant for some viruses (e.g., HIV, [10]), other constraints may be needed in less deadly viral infections [65]. Recent theory posits a role for detection in limiting virulence evolution, where symptoms trigger altered behavior that reduces transmission success [66]. We find no evidence that faster replication hastens symptoms—and therefore associated changes in host behavior—but it could exacerbate symptom severity (morbidity), causing more extreme alterations to host behavior and costly reductions in transmission success following symptom onset. Consistent with that idea, a recent influenza study uncovered a positive correlation between morbidity-induced reductions in host activity and infectiousness [67]. Our within-host modeling suggests that resource limitation may also be an important constraint, and one that has received more attention in recent models of within-host viral dynamics [15]. Theory suggests that resource limitation can generate a tradeoff between transmission early versus late in infection (e.g., in malaria parasites, [68, 69]). If resource limitation imposes a tradeoff between pre- and post-symptomatic transmission, then latency could evolve over the course of an epidemic, with traits that enhance infectiousness later in infection favored as transmission becomes more difficult [70].

One area that warrants further attention is the role of replication rates in modulating the time required for virus to exit the host. For respiratory viruses, the delay between viral replication and transmission would likely be short, given the time required expulsion of viral particles during breathing, coughing, and sneezing. The delay is likely greater for pathogens colonizing the lower respiratory tract, where the site of aerosol formation is farther from the nose or mouth [71]. For enteric pathogens, the lag between replication and fecal shedding is determined by the gut transit time. For simplicity, we assumed no such time lag, but incorporating a delay would not alter our finding that pathogen-based symptom thresholds are inconsistent with the data. Whatever the time required to exit the host, peak pathogen load should always precede peak shedding. If symptoms are assumed to occur when pathogen load crosses a threshold, then peak shedding must necessarily follow symptoms. This result adds to findings in other systems that a within-host abundance threshold by itself is insufficient to explain symptom onset (e.g., the ‘pyrogenic threshold’ of malaria parasite numbers required to trigger malaria fevers, [72]). The implications for other assumptions about symptom onset depend on how viral replication rates influence gut transit times. If a higher replication rate results in shorter gut transit time, that could hasten both shedding and the onset of symptoms like diarrhea, making the impact on pre-symptomatic transmission difficult to predict. Gut transit times are known to vary person-to-person by a factor of days and within individuals day-to-day [73]. Understanding how viral replication rates alter the time required to exit the host would enhance efforts to reconstruct within-host dynamics from CHI data.

We found no significant impact of inoculum dose on viral replication rate in norovirus infections (Fig. S4A). We find a weak trend towards larger doses hastening the time until symptom onset, consistent with previous analysis of this dataset [1], but unlike that study, we find no clear relationship been dose and the timing of peak shedding (Fig. S4B). We attribute this discrepancy to a slight difference in the data fitted, since we truncated time series to avoid fitting values beyond the initial peak and decline of viral abundance, all that can be described by Eq. 1. Comparisons of fits reconstructed using the methods from [1] versus our own reveals that our approach is better able to capture the peak height and timing in norovirus data (Fig. S1). Distinguishing between the end of an infection wave and noise is challenging, especially in the SARS-CoV-2 time series (Fig. S2). Addressing that challenge will be central to understanding how within-host dynamics contribute to the timing of symptoms versus infectiousness. Despite the complexity, general patterns may still emerge. For example, slowed multiplication rates when population sizes are low (known as Allee effects) are thought to apply broadly across diverse organisms [74] and have been documented in pathogenic organisms as diverse as *Vibrio cholerae* [75] and malaria parasites [76]). Applied to CHI trial data, models can reveal whether relationships between dose and pre-symptomatic transmission are idiosyncratic or predictable across different pathogens.

Our simple models allow predictions for how multiple immune parameters—immune effector recruitment and loss rates, and pathogen clearance rate per immune effector—impact pre-symptomatic transmission, enabling comparison with CHI shedding and symptom data [19]. More complex models could predict how specific kinds of immune effectors modulate viral replication, shedding, and symptoms. Data on levels of immune effectors such as cytokines and antibodies are obtainable from CHI trials [44, 77, 78], and models already describe their impact on symptom and viral dynamics (e.g., cytokines, [79]). Coupling these data with model predictions could address a range of open questions, including how vaccination alters the timing of symptom onset versus infectiousness in breakthrough infections, whether immuno-compromised patients are more or less likely to transmit pre-symptomatically, and the extent to which different types of immune effectors promote pre-symptomatic transmission. Accounting for variation in symptom timing due to adaptive immunity will be crucial to predicting disease spread following vaccination and as emerging viruses transition to endemic persistence. If those with immunological memory through previous exposure or vaccinations are more likely to transmit pre-symptomatically, it would be possible to target interventions like testing and masking where they will be most useful.

Understanding the within-host causes of pre-symptomatic transmission is necessary to predict epidemiological impacts. For a given pathogenic organism, variation across individual hosts can have an outsize impact on the potential for outbreaks, with undiagnosed or mis-diagnosed infections implicated in more superspreading events than unusually high contact rates [80]. Our analysis of individual infections reported by [1] and [2] suggests that—even controlling for viral genetic background and dose—there are considerable differences in the propensity for pre-symptomatic transmission (Fig. 3, S6). Individual-level data on the timing of symptoms, coupled with models of symptom onset, can reveal the mechanistic causes of pre-symptomatic transmission and offer insight into the intervention measures best-suited to containing outbreaks.

## 4 Methods

### 4.1 Survey of CHI trials

We searched for viral CHI studies on Google Scholar using the terms: human viral OR load OR shedding OR onset OR symptom “experimental OR controlled infection”. We also searched on PubMed, filtering for English articles using the terms: human AND virus AND (load OR shedding OR onset OR symptom) AND (”experimental infection” OR “challenge study” OR “controlled infection”). The Google Scholar search returned 980 viewable studies (returning an error past page 98 of results), and the PubMed search returned 204 studies. We obtained an additional 30 studies by surveying the references of pertinent reviews, meta-analyses, and secondary analyses found during our searches. We included only human challenge studies published on or after 2003 containing shedding and symptom onset data recorded daily beginning on the day of inoculation, whether those data were reported in tables or graphics. We only considered studies where at least one group of participants did not receive any intervention after inoculation with non-genetically modified viruses that could be classified as enteric or respiratory. The last query performed was on November 6, 2024. Of the 1214 studies screened, we found 65 tracking viral shedding and/or symptom onset, but we excluded 24 studies for reporting only shedding onset or symptom onset, or in which individual symptom scores and shedding could not be distinguished from plots (Table S6). Most studies (56 of 65) presented data as graphs, rather than reporting values in the text.

We recorded mean or median onset (as available) for symptoms and shedding from the text or tables directly when possible. If individual data were reported, we calculated the average onset of shedding and symptoms. If studies reported shedding graphically, we calculated onset as the first day with mean shedding greater than zero. Some studies also reported symptoms as a participant-reported score over time. For those, we estimated the onset of symptoms from the first reported timepoint where the mean/median symptom score was greater than or equal to the minimum possible for a symptomatic individual (usually 1).

We used data from the placebo treatment or control group in case-control studies. Though most studies reported total symptom scores, when scores were separated into categories (e.g., upper versus lower respiratory symptoms, [37]), we used the category with the earliest onset. PCR results for shedding were prioritized over other detection methods. When samples were taken from multiple sites, namely the throat versus nose swabs taken in [2], we used the data leading to the earliest shedding onset.

### 4.2 Identifying correlates of pre-symptomatic transmission

Ge et al. [1] report shedding time series data and incubation periods for 20 participants inoculated with norovirus, where sample timing depended on the availability of fecal samples. Zhou et al. [2] present daily shedding values and symptom scores for 18 participants inoculated with SARS-CoV-2. To fit the four parameters for Eq. 1 to individual shedding trajectories from each study, we minimized the sum squared error (SSE) between observed log_10_ viral shedding and predicted log_10_ viral load using the optim function in **R 4.2.1**. We calculated SSE for 1000 parameter sets chosen by Latin hypercube sampling over plausible bounds and selected the 100 best SSE values for each infection to use as starting guesses for the Nelder-Mead optimization. If any of the 100 optimizations did not converge, we selected the parameter sets with the next lowest SSEs from our initial set of 1000 to use as starting guesses until we obtained 100 fits that converged. Confidence intervals were constructed by resampling with replacement from the 100 optimized parameter fits for each infection with weights 1 *− SSE/SSE_max_*.

Viral abundance often dropped below the limits of detection, and the norovirus data utilized two methods of quantifying abundance with two distinct detection limits [1]. Preliminary fits treating samples below the detection limit as missing generated implausible fits (i.e., predicted abundance far in excess of the detection limit). Instead, for observed values below the detection limit(s), we conditioned the error calculation on whether the model predicted trajectory was above the detection limit, in which case observed values were set equal to the detection limit. When predicted values fell below the lower limit (i.e., 1 or the lower detection limit for the norovirus data), observed abundance was set to the lower limit. When observed and predicted viral abundance fell within the bounds set by detection limits, nothing was added to the SSE calculation. The other complication was that Eq. 1 describes a single peak in infection [20] followed by a log-linear decline, but many infections exhibit multiple peaks (especially the SARS-CoV-2 infections) or a relatively constant level of shedding after a log-linear decline. To account for this variability and obtain robust estimates of replication rates and the timing of peak shedding, we implemented an algorithm to truncate the data to fit. Specifically, we chose as the endpoint whichever of four possibilities that occurred first: (1) the end of the time series; (2) the first value after maximum detected viral abundance that fell below the detection limit and for which viral abundance stayed below detectable values for one day or more; (3) viral abundance below all subsequent values prior to dropping below the detection limit or the end of the time series; (4) a local minimum in viral abundance such that the derivative was negative at two preceding points and positive derivative at two subsequent points. We excluded one participant of the SARS-CoV-2 study who never showed symptoms (Throat 10) from all analyses. Four other participants from that study were excluded from analyses involving model-fitted replication rates or peak shedding timing, for either an unclear peak (Throat 2), or for having fewer than 5 estimates of viral abundance within the fitted data (Throat 1, Throat 6, Throat 14, and Throat 15). We find that our approach better captures the timing and magnitude of peak viral shedding than the Bayesian methods employed previously to fit the entire time series [1] (Fig. S1).

### 4.3 Defining a plausible within-host model for pre-symptomatic transmission

We extended the within-host model described in [22] to incorporate thresholds for symptom onset once the abundance of either pathogen (*P*) or immune effectors (*X*) exceeds a threshold (denoted *s_P_* or *s_X_* respectively). This “base model” assumes exponential pathogen replication at rate *r* that is limited by immune clearance:

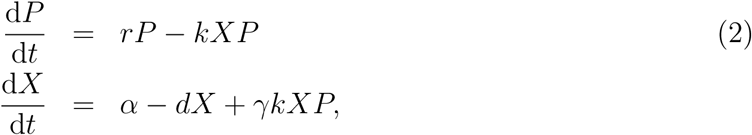

where *k* indicates the rate of immune removal given contact with pathogen, *α* and *d* denote the baseline rates of immune effector proliferation and removal (respectively), and *γ* represents immune effector proliferation due to contact with pathogen. We then modify this model to incorporate immune inhibition of pathogen replication instead of direct clearance, following innate immune dynamics from [81]:

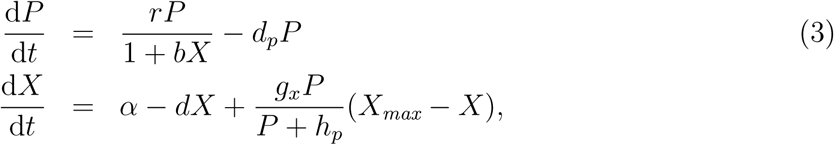

where *b* scales the impact of immune effectors on pathogen replication, *d_p_* indicates the background rate of pathogen mortality, *g_x_* determines how quickly immune effectors proliferate (or are recruited) as pathogen numbers increase, a response that saturates with large numbers of pathogens according to the constant *h_p_*, and *X_max_* specifies the maximum upregulation of immunity. Finally, we reformulate model 2 to relax the assumption of constant, exponential replication by imposing a carrying capacity while retaining the base model immune dynamics:

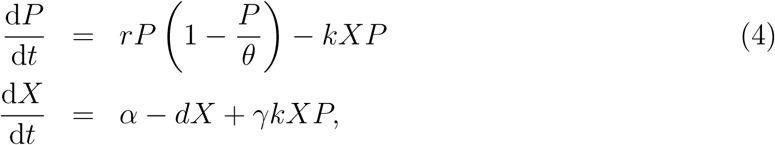

where *θ* represents the carrying capacity. For each of models 2-4, we simulated dynamics while varying replication rates (*r*) to investigate the consequences for the relative timing of peak pathogen load relative to symptom onset. For simplicity, we assume that viral shedding is equivalent to viral load within the host with no time delay. Parameter values are listed in Table S8.

#### Analysis of peak timing

Our simulations suggest that symptom onset follows peak shedding at low replication rates in systems of equations 2 and 3 and at high replication rates (the empirical pattern) in model 4. To corroborate these patterns, we solved for peak pathogen abundance by setting d*P/*d*t* = 0. Whether that condition holds is solely a function of immune effector abundance (*X*) in systems of equations 2 and 3 (*X* = *r/k* and *X* = (*r − d_p_*)*/bd_p_*, respectively). Thus, as replication rates (*r*) increase, immune effector abundance (*X*) must attain higher levels to force pathogen numbers to peak and then decline. For a given symptom threshold *s_x_*, it is more likely that immune effector abundance will cross that threshold prior to the peak in pathogen abundance as replication rates (*r*) increase. Hence pre-symptomatic transmission is most likely at low replication rates, in opposition to the patterns observed in CHI data. When pathogen dynamics are subject to a carrying capacity, d*P/*d*t* = 0 when *X* = *r*(1*−P/θ*)*/k*, so that the timing of the peak is not a function of immune effector abundance alone. Since the timing of the peak (*dP/dt* = 0) is conditioned on another state variable (here *P*), immune effector abundance *X* need not attain a larger value to enforce a pathogen peak at higher replication rates, creating the possibility of hastening peak shedding with minimal impact on symptom onset.

## Data Availability

Full code to reproduce analyses and figures is available at \\
\href{https://github.com/greischarlab/presymptomatic-transmission}{github.com/greischarlab/pre-symptomatic-transmission}. The norovirus data published by \cite{Ge2023} and associated analysis can be found at \href{https://github.com/yangepigroup/eid230117}{github.com/yangepigroup/eid230117}, save for the sample weights that were kindly provided by Dr. Andreas Handel. Data from the SARS-CoV-2 study \cite{Zhou2023} are available by request from the study authors.

https://github.com/greischarlab/presymptomatic-transmission

The authors declare no competing interest

## Data availability

Full code to reproduce analyses and figures is available at github.com/greischarlab/pre-symptomatic-transmission. The norovirus data published by [1] and associated analysis can be found at github.com/yangepigroup/eid230117, save for the sample weights that were kindly provided by Dr. Andreas Handel. Data from the SARS-CoV-2 study [2] are available by request from the study authors.

## Acknowledgements.

We thank Chadi Saad-Roy and Nicole Mideo for helpful discussion, Louis Gross for insightful questions, and Andreas Handel for invaluable criticism of an earlier version of this study. We greatly appreciate Drs. Wendy Barclay and Jie Zhou sharing SARS-CoV-2 data from their recent study. This work was supported by the Cornell University College of Agriculture and Life Sciences (M.A.G.), and the Cornell Institute of Host-Microbe Interactions & Disease Undergraduate Research Experience (K.Z.).

## Supplemental figures & tables

**Figure S1:**
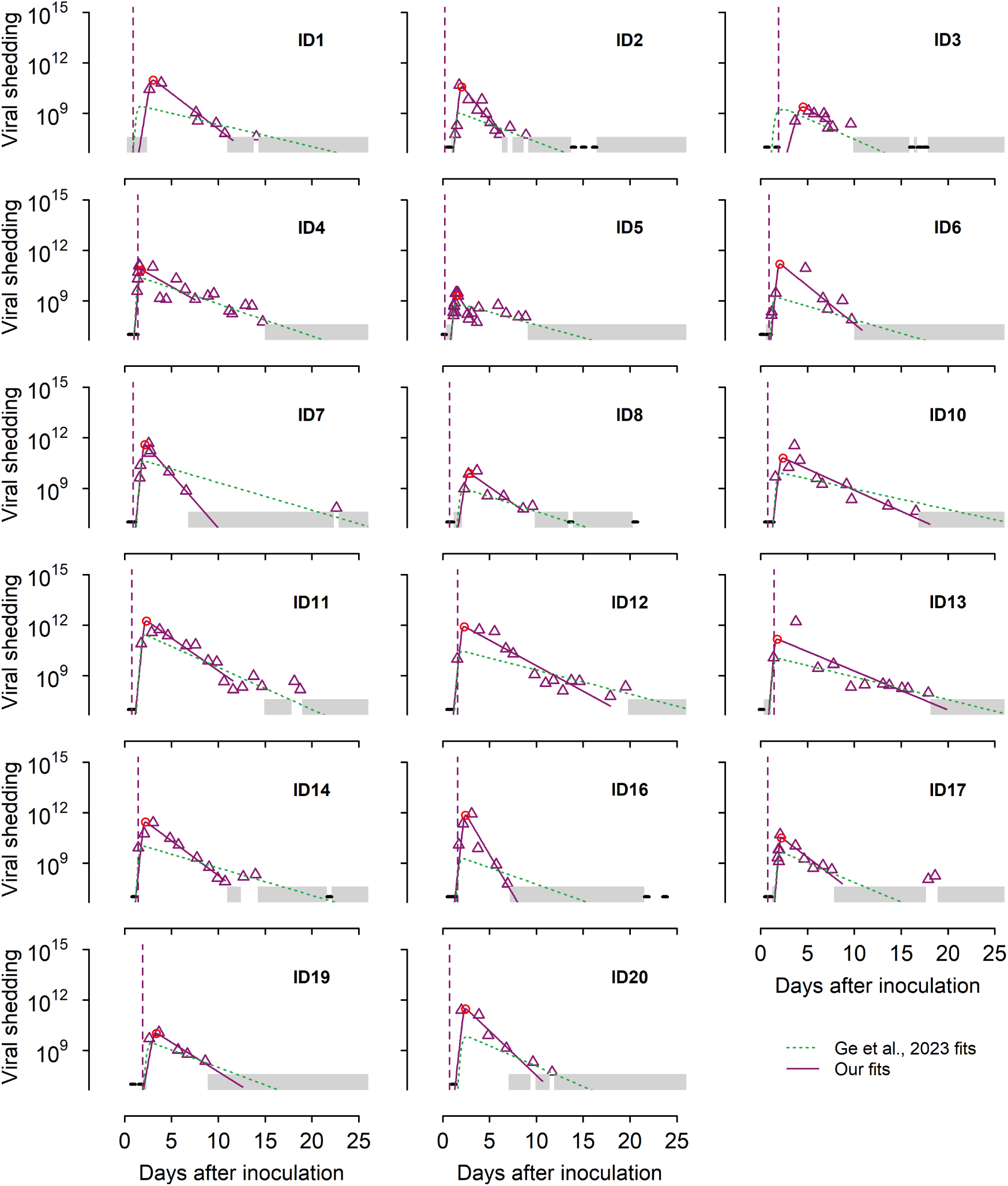
No participants demonstrate pre-symptomatic transmission in the norovirus study [1]. The triangles indicate viral shedding, the solid purple line indicates the fit of Equation 1, and the dotted vertical lines indicate the onset of symptoms. A gray rectangle indicates the range of shedding values for samples that fall below the detection limit for the quantitative qRT-PCR method (4 *×* 10^7^ genome equivalent copies / g of stool) but above the limit for the qualitative IMC RT-PCR method (15,000 genome equivalent copies / g of stool). Horizontal black lines indicate samples that fall below both detection limits. The ID number of each participant is labeled in each plot. The dotted green lines indicate fits using the Bayesian methods described by [1].

**Figure S2:**
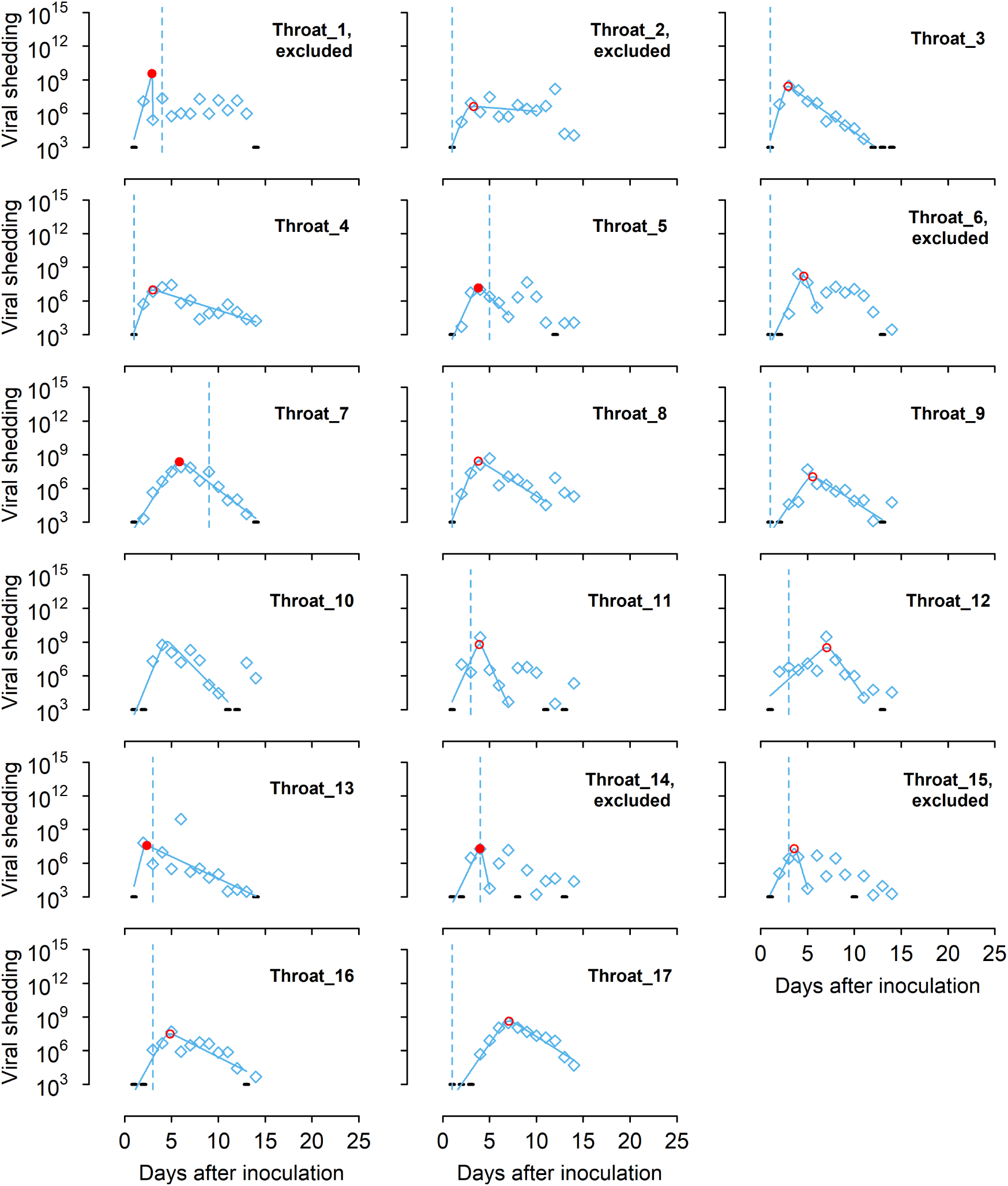
Five participants demonstrate pre-symptomatic transmission in the SARS-CoV-2 study [2]. The diamonds indicate viral shedding, the solid line indicates the fit of Equation 1, and the dotted vertical lines indicate the onset of symptoms. Black lines indicate samples where no virus was detected. Infection ID Throat 2 was excluded due to an unclear peak, and Throat 1, Throat 6, Throat 14, and Throat 15 were excluded due to having fewer than 5 observations during the first wave of infection (see Methods for details on determining the end point).

**Figure S3:**
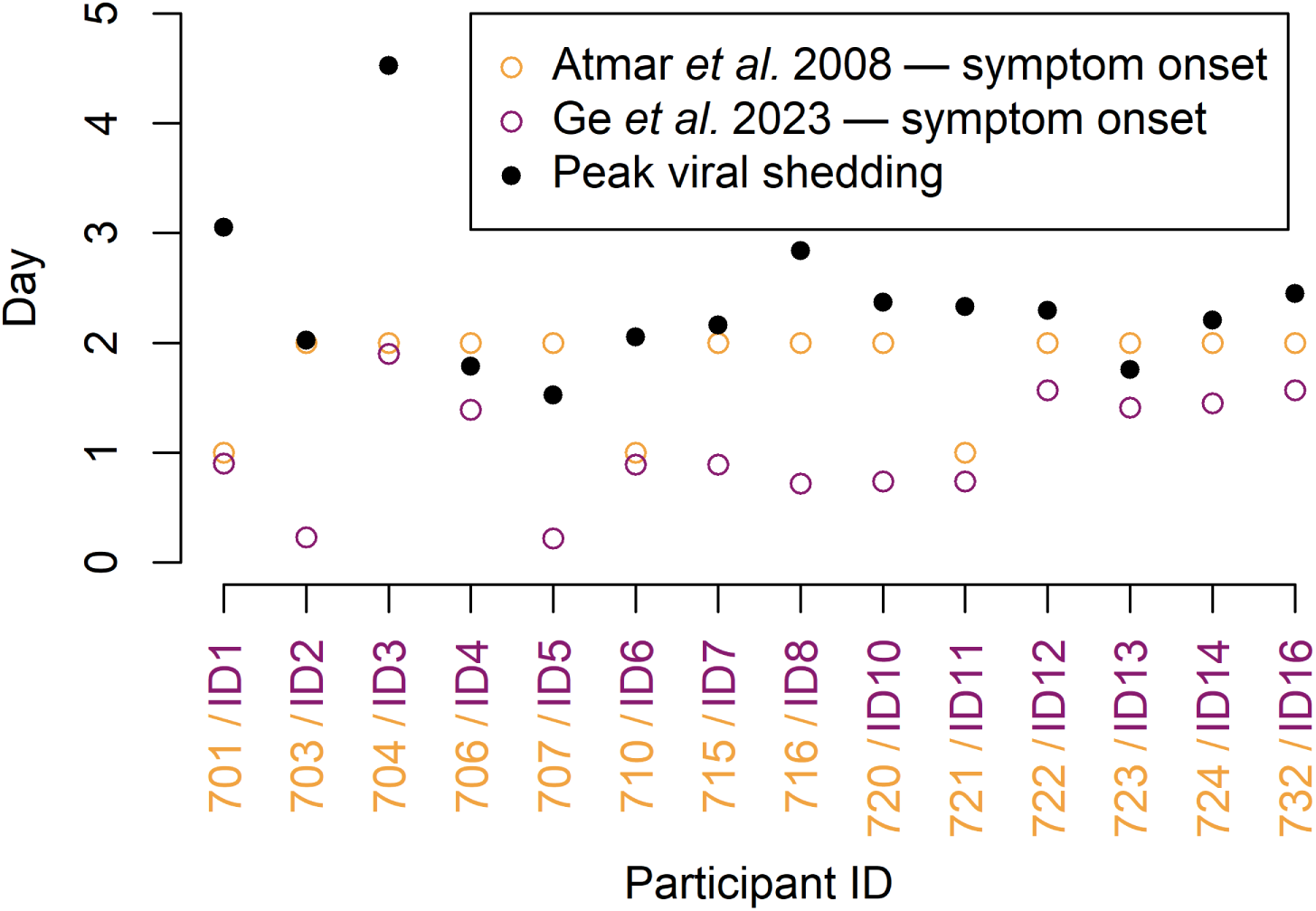
The symptom onset times reported by [19] (orange open circles) differ from those recorded in the reanalysis performed in [1] (open purple circles). Most but not all of the differences are explained by the fact that [19] report in units of whole days, while [1] provide higher resolution. Closed black dots indicate our estimates for the time of peak viral shedding, shown for comparison.

**Figure S4:**
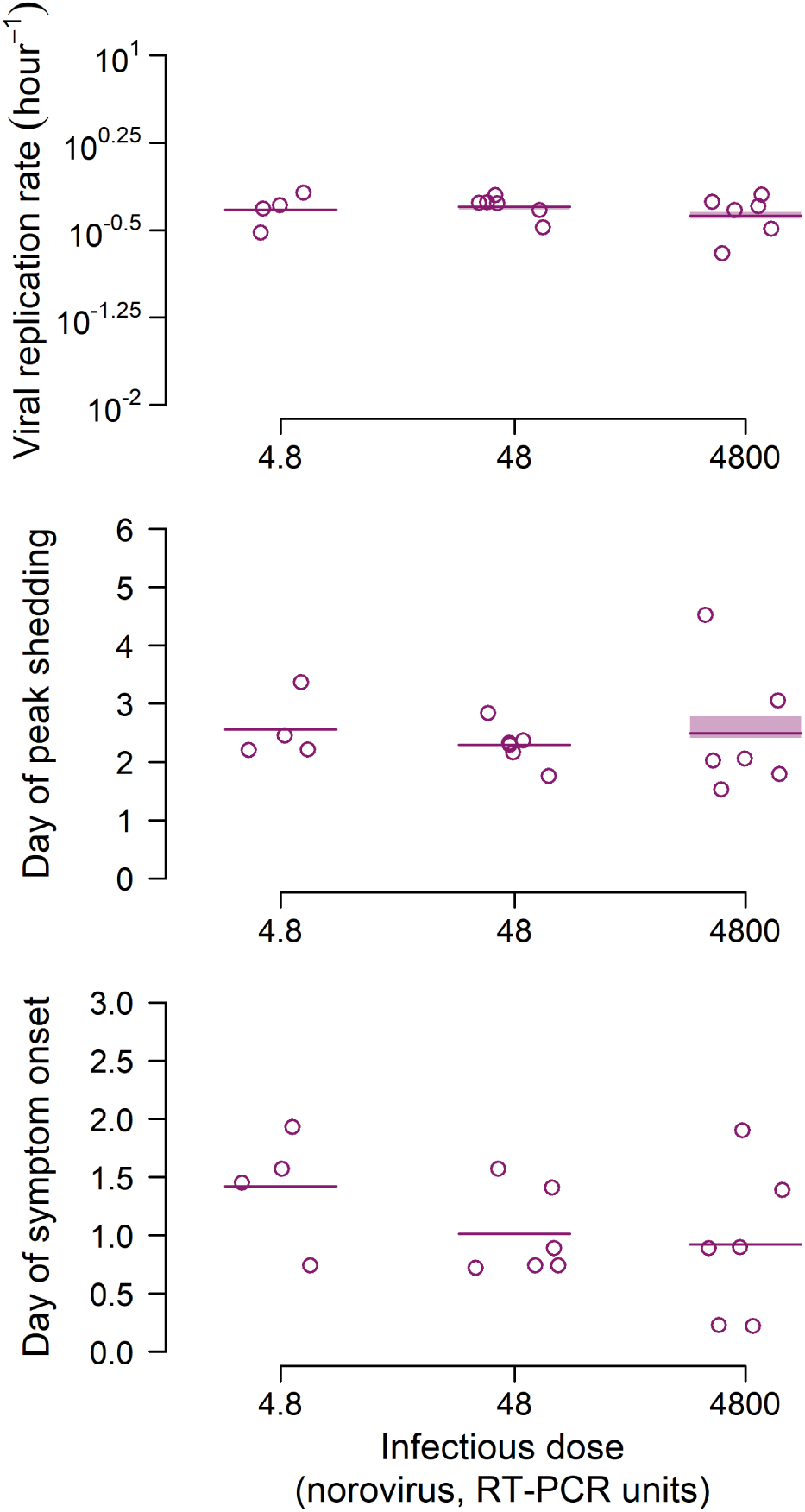
For participants in the norovirus study [1], inoculum dose has no impact on viral replication rate or time of peak viral load, but it does demonstrate a nonsignificant negative correlation with the time of symptom onset. Horizontal lines indicate the mean for each dose. Data was excluded for the lowest dose (0.48 RT-PCR units) because it contained only one individual. Shaded regions show 95% confidence intervals calculated from bootstrapping (see Methods for details). Panel C has no confidence interval because symptom onset was observed rather than fitted.

**Figure S5:**
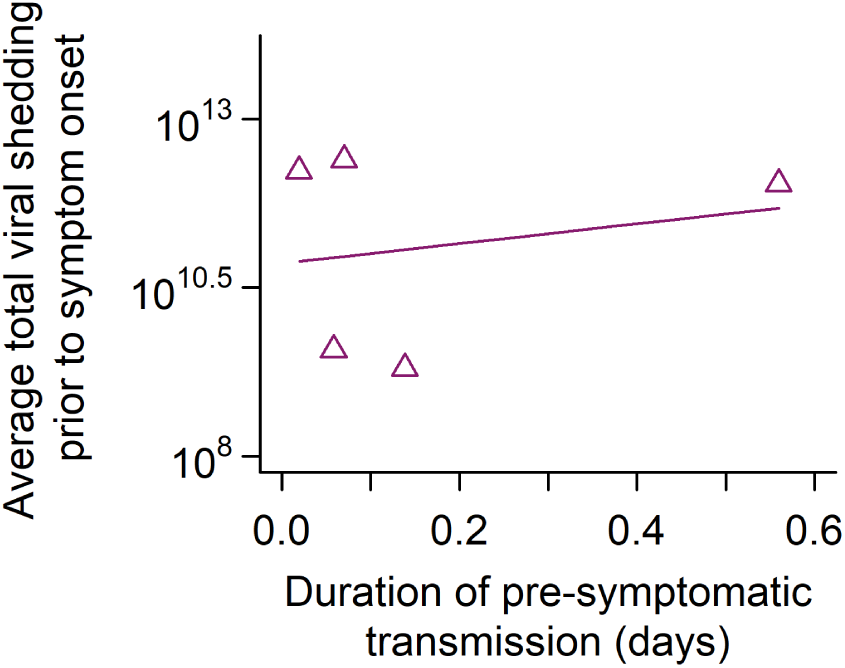
There is no correlation between the average amount of virus shed in the pre-symptomatic period and the duration of that pre-symptomatic period for participants in the norovirus study [1]. Amount of virus shed was calculated by multiplying the concentration of virus at each time point by the sample weight. Multiple estimates for the same time point were averaged prior to averaging across time points, as we assume these are multiple measurement attempts for the same sample.

**Figure S6:**
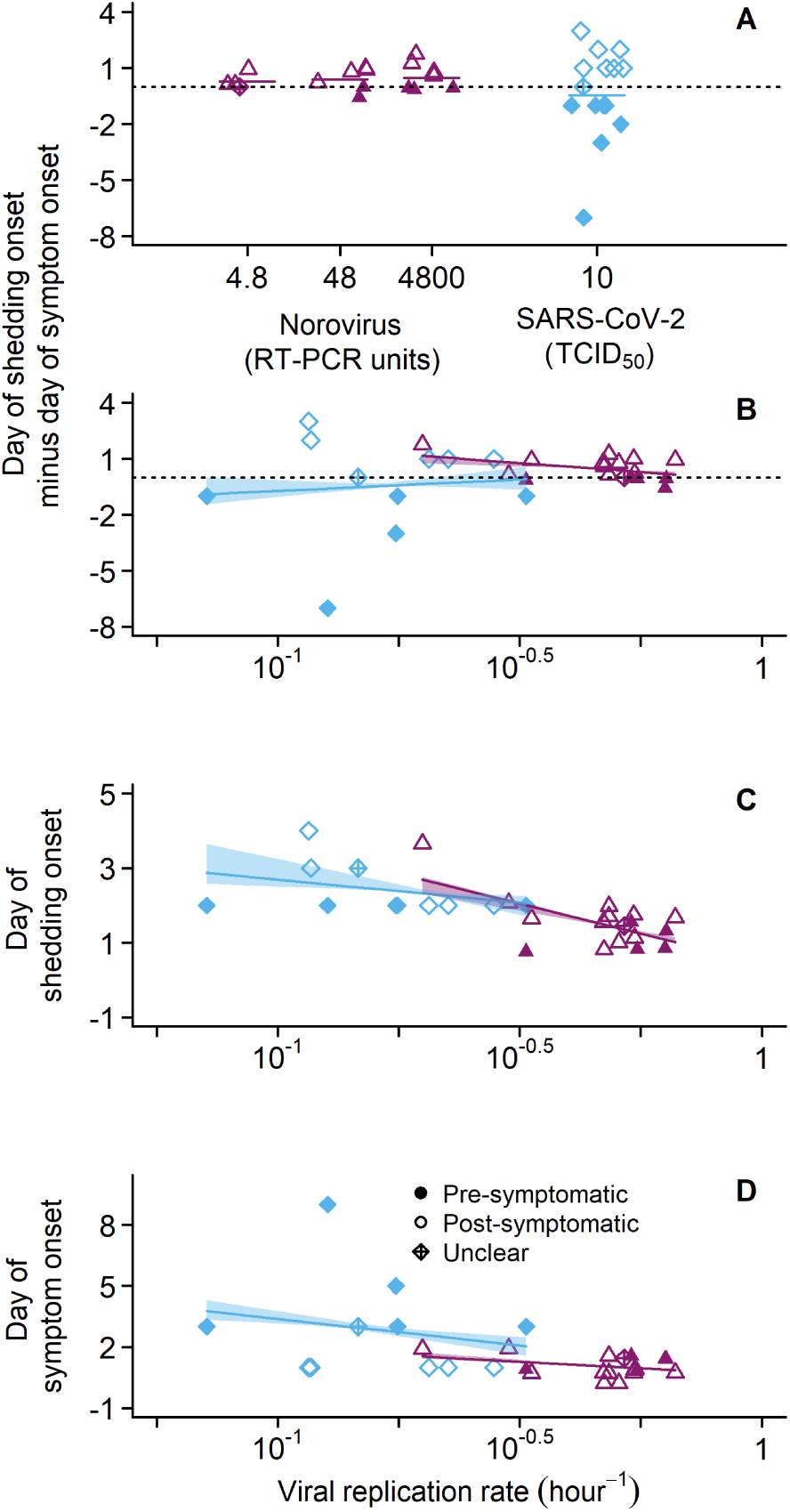
The impact of dose and replication rates is less clear when pre-symptomatic transmission is defined less conservatively as shedding preceding symptom onset. (A) Norovirus [1] (purple triangles) and SARS-CoV-2 [2] (blue diamonds) show the delay between symptom and shedding onset for each participant, with horizontal bars to indicate the average delay for each pathogen. Closed symbols indicate shedding began prior to symptom onset, with one SARS-CoV-2 participant exhibiting apparent simultaneous onset of shedding and symptoms (open diamond with cross), making it unclear whether symptoms or shedding began first. (B) The delay between symptom onset and shedding onset does not change with faster estimated viral replication rate (*P*_2_, Eq. 1). (C) There are negative correlations between replication rate and shedding onset (significant for norovirus, *p* = 0.0061, *R*^2^ = 0.40, nonsignificant for SARS-CoV-2, *p* = 0.28, *R*^2^ = 0.12). Panel D is identical to Fig. 3D and placed here for ease of comparison. Shaded regions show 95% confidence intervals calculated by bootstrapping from fits for each participant (see Methods for details).

**Figure S7:**
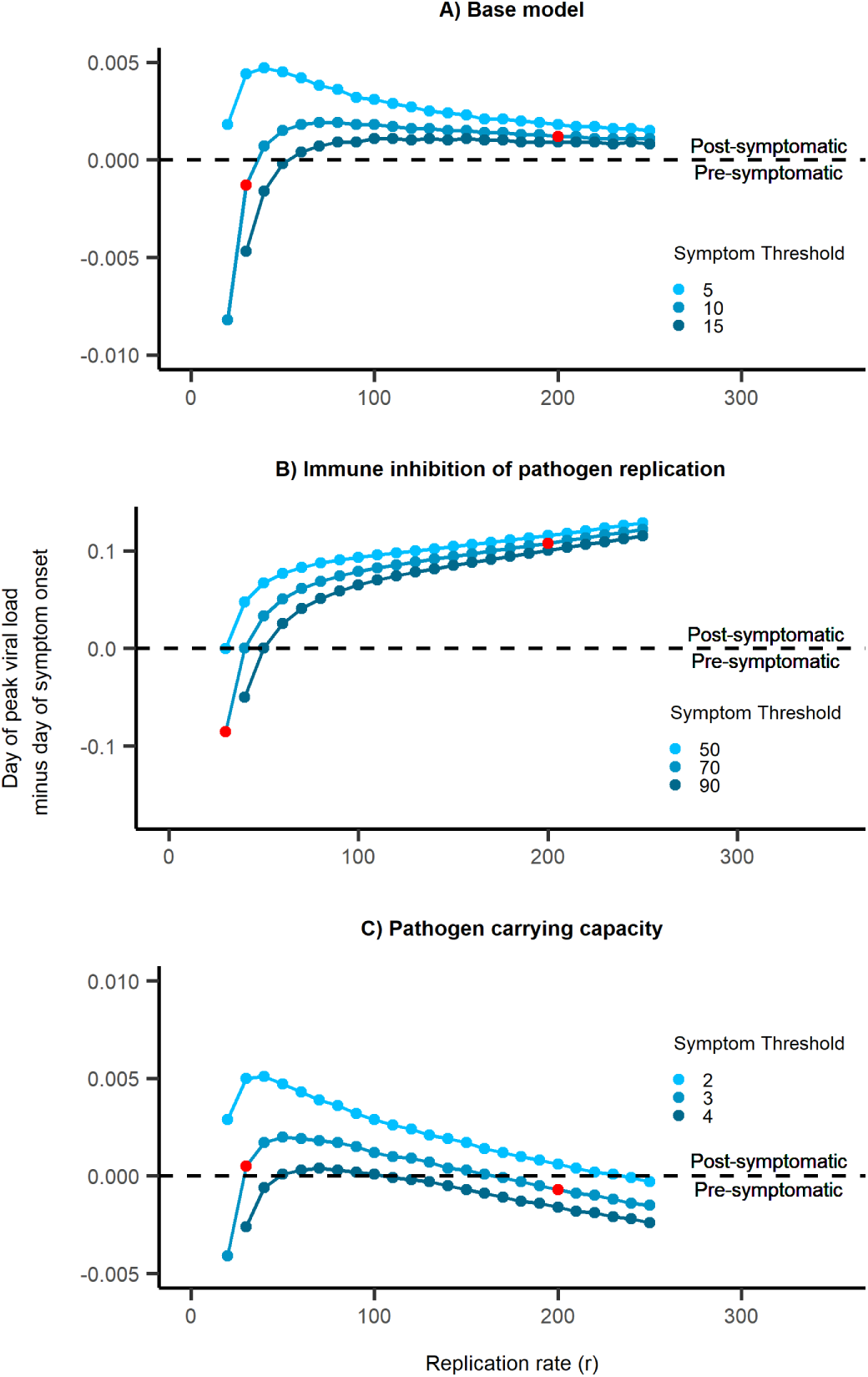
In simulations, the relative timing of symptom onset and peak shedding varies with replication rates. All models shown here assume that symptom onset occurs when immune effector abundance exceeds an arbitrary threshold *s_X_*. In the base model (Eqns. 2), a higher pathogen replication rate (*r*) decreases the chance of pre-symptomatic transmission (A). When we assume that immunity interferes with pathogen replication rather than direct removal and that immune upregulation saturates at a certain level (Eqns. 3, *X_max_* = 5 *×* 10^15^), then faster replication reduces pre-symptomatic transmission (B). If we instead impose a carrying capacity (*θ* = 5 *×* 10^5^) while assuming direct removal of pathogens (Eqns. 4, *k*=7), faster replication first reduces then increases pre-symptomatic transmission (C). Symptom thresholds *s_X_* from low to high are 5, 10, and 15 for panel A; and 50, 70, 90 for panel B; and 2, 3, 4 for panel C. The replication rates used in Fig. 4B, C, and D are highlighted in red.

**Figure S8:**
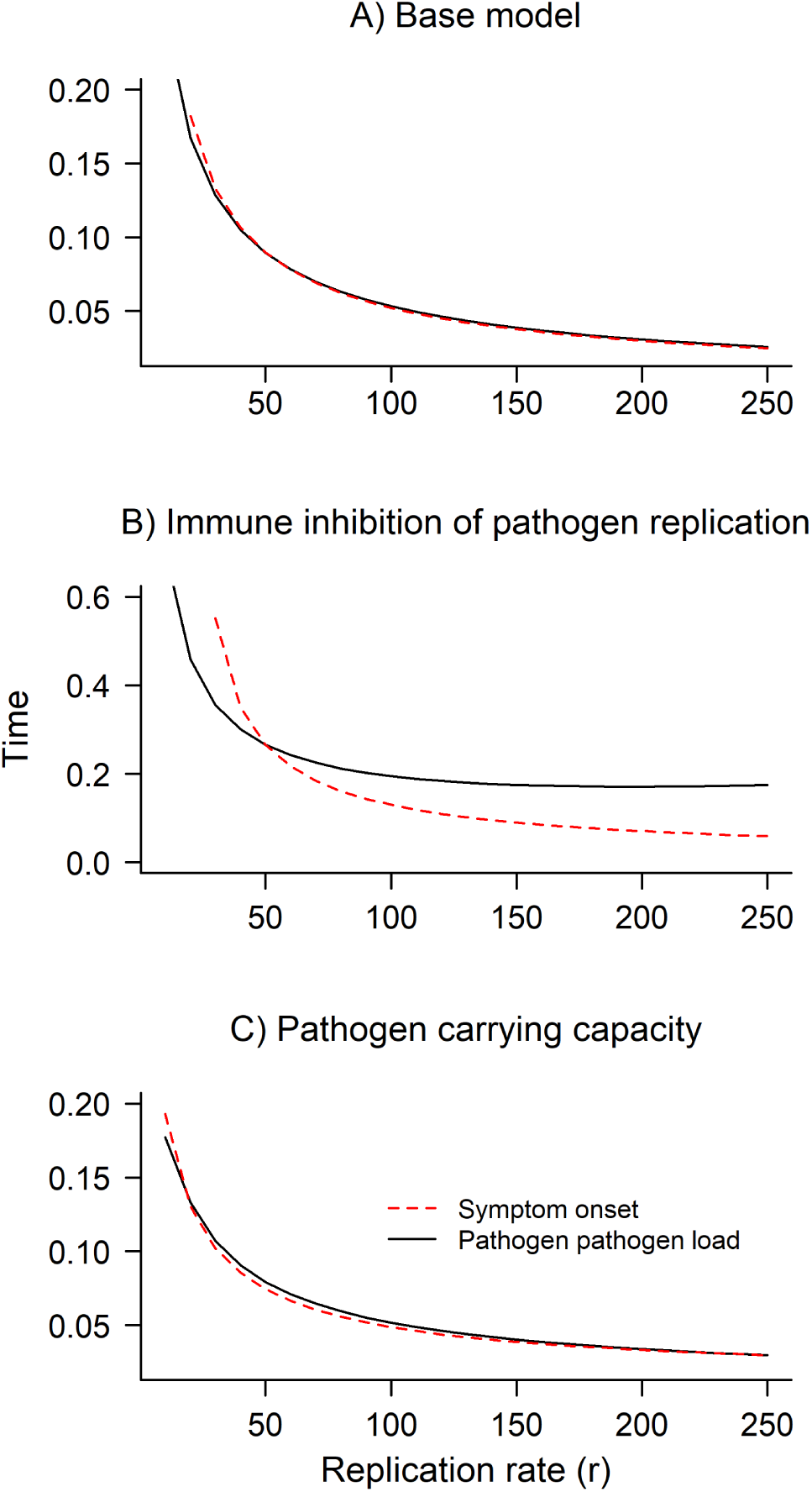
With the assumption of exponential pathogen growth in our base model (Eqns. 2), we see that at low replication rates, peak pathogen load occurs before symptom onset, and that at high replication rates, it occurs after symptom onset (A). A similar pattern emerges if we assume that the immune response is inhibiting pathogen replication rather than directly killing pathogens (Eqns. 3) (B). In contrast, if we assume that there is a pathogen carrying capacity with direct killing of pathogens (Eqns. 4), then at very low replication rates, peak pathogen load occurs before symptom onset; at intermediate replication rates, peak pathogen load occurs after symptom onset; and at high replication rates, peak pathogen load again occurs before symptom onset (C).

**Figure S9:**
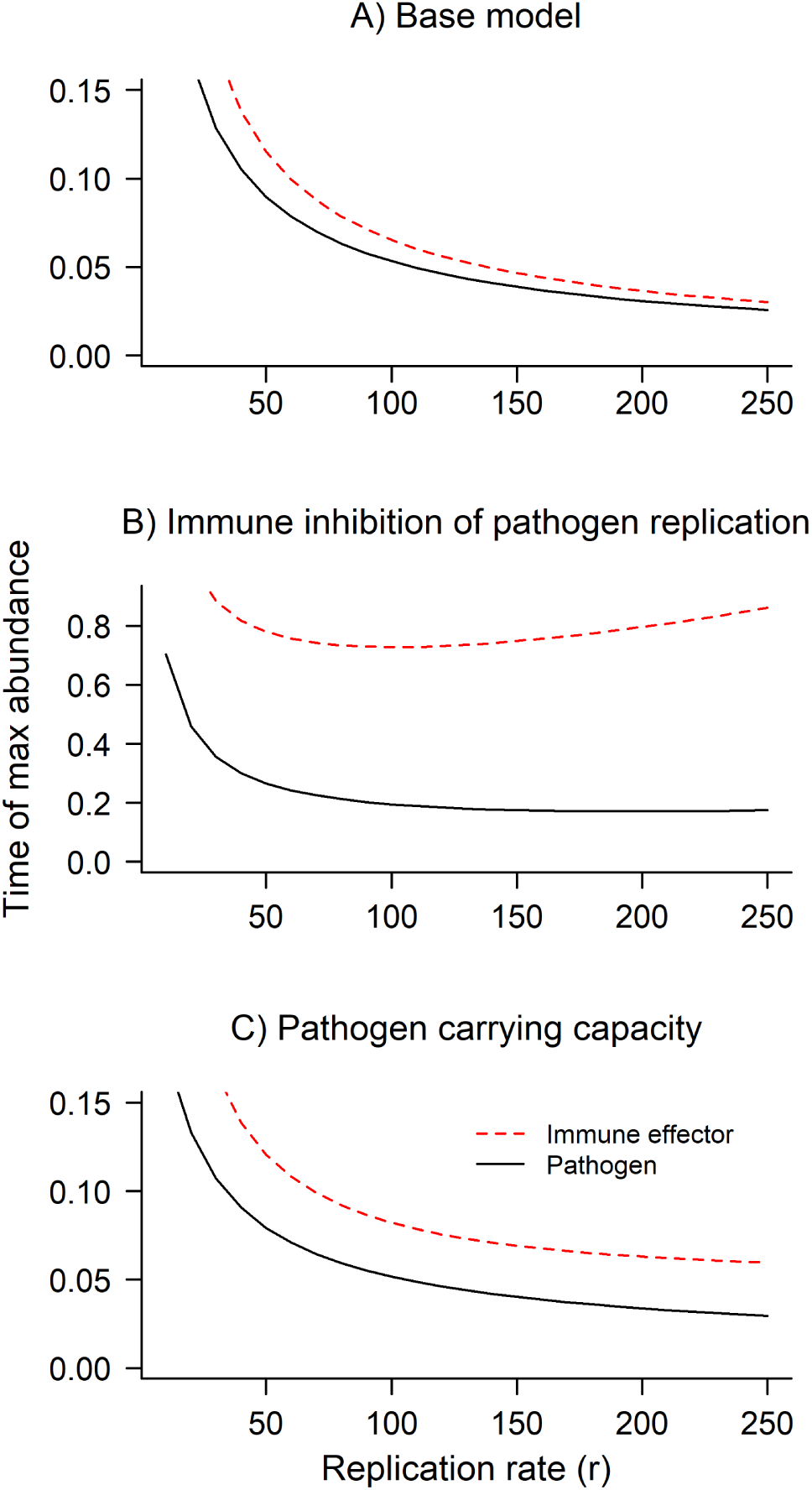
Immune response (X) peaks after pathogen load (P) in all three of our models (Eqns. 2-4) for all of the pathogen replication rates we used (*r* = 10 to 250 by increments of 10).

**Table S1:** rhinovirus studies.

| Table S1: rhinovirus studies | Average onset (days) |  |
| --- | --- | --- |
|  | Shedding | Symptoms |
| E. Grzesiak et al. Assessment of the Feasibility of Using Non-invasive Wearable Biometric Monitoring Sensors to Detect Influenza and the Common Cold Before Symptom Onset. *JAMA Network Open* 4 (2021), e2128534. DOI: 10.1001/jamanetworkopen.2021.28534 | 1.3 | 0.8 |
| D. J. Jackson et al. IL-33-Dependent Type 2 Inflammation during Rhinovirus-induced Asthma Exacerbations *in vivo*. *American Journal of Respiratory and Critical Care Medicine* 190 (2014), 1373–1382. DOI: 10.1164/rccm.201406-10390C | 1.7 | 2 |
| F. G. Hayden et al. Phase II, Randomized, Double-Blind, Placebo-Controlled Studies of Rupintrivir Nasal Spray 2-Percent Suspension for Prevention and Treatment of Experimentally Induced Rhinovirus Colds in Healthy Volunteers. *Antimicrobial Agents and Chemotherapy* 47 (2003), 3907–3916. DOI: 10.1128/AAC.47.12.3907-3916.2003 | 1 | 1 |
| D. J. Fullen et al. A Tool for Investigating Asthma and COPD Exacerbations: a Newly Manufactured and Well Characterised GMP Wild-Type Human Rhinovirus for Use in the Human Viral Challenge Model. *PLOS ONE* 11 (2016), e0166113. DOI: 10.1371/journal.pone.0166113 | 2.13 | 2.5 |
| K. M. Peterson et al. Effects of dietary supplementation with conjugated linoleic acid on experimental human rhinovirus infection and illness. *Antiviral Therapy* 14 (2009), 33–43. DOI: 10.1177/135965350901400111 | 1 | 1 |
| P. Mallia et al. An experimental model of rhinovirus induced chronic obstructive pulmonary disease exacerbations: a pilot study. *Respiratory Research* 7 (2006), 116. DOI: 10.1186/1465-9921-7-116 | 2 | 3 |
| J. C. L. Spurrell et al. Human airway epithelial cells produce IP-10 (CXCL10) in vitro and in vivo upon rhinovirus infection. *American Journal of Physiology-Lung Cellular and Molecular Physiology* 289 (2005), L85–L95. DOI: 10.1152/ajplung.00397.2004 | 1 | 1 |
| J. P. DeMore et al. Similar colds in subjects with allergic asthma and nonatopic subjects after inoculation with rhinovirus-16. *Journal of Allergy and Clinical Immunology* 124 (2009), 245–252.e3. DOI: 10.1016/j.jaci.2009.05.030 | 1 | 0 |

| Table S1 continued: rhinovirus studies | Average onset (days) |  |
| --- | --- | --- |
|  | Shedding | Symptoms |
| P. Mallia et al. Rhinovirus Infection Induces Degradation of Antimicrobial Peptides and Secondary Bacterial Infection in Chronic Obstructive Pulmonary Disease. *American Journal of Respiratory and Critical Care Medicine* 186 (2012), 1117–1124. DOI: 10.1164/rccm.201205-08060C | 5 | 3 |
| S. D. Message et al. Rhinovirus-induced lower respiratory illness is increased in asthma and related to virus load and Th1/2 cytokine and IL-10 production. *Proceedings of the National Academy of Sciences* 105 (2008), 13562–13567. DOI: 10.1073/pnas.0804181105 | 2 | 6 |
| R. Lutter et al. The Dietary Intake of Carrot-Derived Rhamnogalacturonan-I Accelerates and Augments the Innate Immune and Anti-Viral Interferon Response to Rhinovirus Infection and Reduces Duration and Severity of Symptoms in Humans in a Randomized Trial. *Nutrients* 13 (2021), 4395. DOI: 10.3390/nu13124395 | 3 | 1 |
| P. Mallia et al. Experimental Rhinovirus Infection as a Human Model of Chronic Obstructive Pulmonary Disease Exacerbation. *American Journal of Respiratory and Critical Care Medicine* 183 (2011), 734–742. DOI: 10.1164/rccm.201006-08330C | 1 | 1 |

**Table S2:** influenza A studies.

| Table S2: influenza A studies | Average onset (days) |  |
| --- | --- | --- |
|  | Shedding | Symptoms |
| J. Eiden et al. M2-Deficient Single-Replication Influenza Vaccine-Induced Immune Responses Associated With Protection Against Human Challenge With Highly Drifted H3N2 Influenza Strain. *The Journal of Infectious Diseases* 226 (2022), 83–90. DOI: 10.1093/infdis/jiab374 | 1.44 | 2 |
| E. Grzesiak et al. Assessment of the Feasibility of Using Non-invasive Wearable Biometric Monitoring Sensors to Detect Influenza and the Common Cold Before Symptom Onset. *JAMA Network Open* 4 (2021), e2128534. DOI: 10.1001/jamanetworkopen.2021.28534 | 2.58 | 2.58 |
| S. E. Sloan et al. Clinical and virological responses to a broad-spectrum human monoclonal antibody in an influenza virus challenge study. *Antiviral Research* 184 (2020), 104763. DOI: 10.1016/j.antiviral.2020.104763 | 0.4 | 1 |

| Table S2 continued: influenza A studies | Average onset (days) |  |
| --- | --- | --- |
|  | Shedding | Symptoms |
| P. J. Lillie et al. Preliminary Assessment of the Efficacy of a T-Cell-Based Influenza Vaccine, MVA-NP+M1, in Humans. *Clinical Infectious Diseases* 55 (2012), 19–25. DOI: 10.1093/cid/cis327 | 1.8 | 1.56 |
| E. L. Ramos et al. Efficacy and Safety of Treatment With an Anti-M2e Monoclonal Antibody in Experimental Human Influenza. *Journal of Infectious Diseases* 211 (2015), 1038–1044. DOI: 10.1093/infdis/jiu539 | 1 | 2 |
| T. M. Wilkinson et al. Preexisting influenza-specific CD4+ T cells correlate with disease protection against influenza challenge in humans. *Nature Medicine* 18 (2012), 274–280. DOI: 10.1038/nm.2612 | 1 | 1 |
| M. J. Memoli et al. Evaluation of Antihemagglutinin and Antineuraminidase Antibodies as Correlates of Protection in an Influenza A/H1N1 Virus Healthy Human Challenge Model. *mBio* 7 (2016). DOI: 10.1128/mBio.00417-16 | 1 | 1 |
| M. J. Memoli et al. Influenza A Reinfection in Sequential Human Challenge: implications for Protective Immunity and “Universal” Vaccine Development. *Clinical Infectious Diseases* 70 (2020), 748–753. DOI: 10.1093/cid/ciz281 | 1 | 1.5 |
| M. J. Memoli et al. Validation of the Wild-type Influenza A Human Challenge Model H1N1pdMIST: an A(H1N1)pdm09 Dose-Finding Investigational New Drug Study. *Clinical Infectious Diseases* 60 (2015), 693–702. DOI: 10.1093/cid/ciu924 | 2 | 3 |

**Table S3:** SARS-CoV-2 studies.

| Table S3: SARS-CoV-2 studies | Average onset (days) |  |
| --- | --- | --- |
|  | Shedding | Symptoms |
| B. Killingley et al. Safety, tolerability and viral kinetics during SARS-CoV-2 human challenge in young adults. *Nature Medicine* 28 (2022), 1031–1041. DOI: 10.1038/s41591-022-01780-9 | 1.67 | 2 |
| J. Zhou et al. Viral emissions into the air and environment after SARS-CoV-2 human challenge: a phase 1, open label, first-in-human study. *The Lancet Microbe* 4 (2023), e579–e590. DOI: 10.1016/S2666-5247(23)00101-5 | 2.41 | 2.75 |

**Table S4:** RSV studies.

| Table S4: RSV studies | Average onset (days) |  |
| --- | --- | --- |
|  | Shedding | Symptoms |
| B. Bagga et al. Unrecognized prolonged viral replication in the pathogenesis of human RSV infection. *Journal of Clinical Virology* 106 (2018), 1–6. DOI: 10.1016/j.jcv.2018.06.014 | 2 | 4 |
| G. Kelly et al. Use of qualitative integrative cyclor PCR (qicPCR) to identify optimal therapeutic dosing time-points in a Respiratory Syncytial Virus Human Viral Challenge Model (hVCM). *Journal of Virological Methods* 224 (2015), 83–90. DOI: 10.1016/j.jviromet.2015.08.019 | 3 | 1.7 |
| J. DeVincenzo et al. Safety and Antiviral Effects of Nebulized PC786 in a Respiratory Syncytial Virus Challenge Study. *The Journal of Infectious Diseases* 225 (2022), 2087–2096. DOI: 10.1093/infdis/jiaa716 | 0.5 | 0.67 |
| B. Schmoele-Thoma et al. Vaccine Efficacy in Adults in a Respiratory Syncytial Virus Challenge Study. *New England Journal of Medicine* 386 (2022), 2377–2386. DOI: 10.1056/NEJMoA2116154 | 3.3 | 5 |
| J. P. DeVincenzo et al. Activity of Oral ALS-008176 in a Respiratory Syncytial Virus Challenge Study. *New England Journal of Medicine* 373 (2015), 2048–2058. DOI: 10.1056/NEJMoA1413275 | 0.5 | 0.67 |
| J. P. DeVincenzo et al. Oral GS-5806 Activity in a Respiratory Syncytial Virus Challenge Study. *New England Journal of Medicine* 371 (2014), 711–722. DOI: 10.1056/NEJMoA1401184 | 2.8 | 1.5 |
| M. S. Habibi et al. Neutrophilic inflammation in the respiratory mucosa predisposes to RSV infection. *Science* 370 (2020). DOI: 10.1126/science.aba9301 | 4 | 3 |
| J. Sadoff et al. Prevention of Respiratory Syncytial Virus Infection in Healthy Adults by a Single Immunization of Ad26.RSV.preF in a Human Challenge Study. *The Journal of Infectious Diseases* 226 (2022), 396–406. DOI: 10.1093/infdis/jiab003 | 2 | 2.9 |
| A. Guvenel et al. Epitope-specific airway-resident CD4+ T cell dynamics during experimental human RSV infection. *Journal of Clinical Investigation* 130 (2019), 523–538. DOI: 10.1172/JCI131696 | 3 | 4 |

| Table S4 continued: RSV studies | Average onset (days) |  |
| --- | --- | --- |
|  | Shedding | Symptoms |
| J. DeVincenzo et al. A randomized, double-blind, placebo-controlled study of an RNAi-based therapy directed against respiratory syncytial virus. *Proceedings of the National Academy of Sciences* 107 (2010), 8800–8805. DOI: 10.1073/pnas.0912186107 | 3.5 | 3 |
| E. Jordan et al. Reduced Respiratory Syncytial Virus Load, Symptoms, and Infections: a Human Challenge Trial of MVA-BN-RSV Vaccine. *The Journal of Infectious Diseases* 228 (2023), 999–1011. DOI: 10.1093/infdis/jiad108 | 2 | 4.5 |
| A. Jozwik et al. RSV-specific airway resident memory CD8+ T cells and differential disease severity after experimental human infection. *Nature Communications* 6 (2015), 10224. DOI: 10.1038/ncomms10224 | 1 | 5 |
| F. Lee et al. Experimental infection of humans with A2 respiratory syncytial virus. *Antiviral Research* 63 (2004), 191–196. DOI: 10.1016/j.antiviral.2004.04.005 | 3 | 1 |
| J. DeVincenzo et al. A randomized, placebo-controlled, respiratory syncytial virus human challenge study of the antiviral efficacy, safety, and pharmacokinetics of RV521, an inhibitor of the RSV-F protein. *Antimicrobial Agents and Chemotherapy* 64 (2020). DOI: 10.1128/aac.01884-19 | 0.5 | 0.5 |
| J. P. DeVincenzo et al. Viral load drives disease in humans experimentally infected with respiratory syncytial virus. *American Journal of Respiratory and Critical Care Medicine* 182 (2010), 1305–1314. DOI: 10.1164/rccm.201002-02210C | 3.1 | 4 |

**Table S5:** norovirus studies.

| Table S5: norovirus studies | Average onset (days) |  |
| --- | --- | --- |
|  | Shedding | Symptoms |
| J. S. Leon et al. Randomized, Double-Blinded Clinical Trial for Human Norovirus Inactivation in Oysters by High Hydrostatic Pressure Processing. *Applied and Environmental Microbiology* 77 (2011), 5476–5482. DOI: 10.1128/AEM.02801-10 | 3.13 | 2.75 |
| Y. Ge et al. Effect of Norovirus Inoculum Dose on Virus Kinetics, Shedding, and Symptoms. *Emerging Infectious Diseases* 29.7 (2023), 1349–1356. DOI: 10.3201/eid2907.230117 | 1.52 | 1.06 |

| Table S5 continued: norovirus studies | Average onset (days) |  |
| --- | --- | --- |
|  | Shedding | Symptoms |
| D. I. Bernstein et al. Norovirus vaccine against experimental human G11.4 virus illness: a challenge study in healthy adults. *The Journal of Infectious Diseases* 211 (2015), 870–878. DOI: 10.1093/infdis/jiu497 | 1 | 1.38 |
| A. E. Kirby et al. Disease course and viral shedding in experimental Norwalk virus and Snow Mountain virus infection. *Journal of Medical Virology* 86 (2014), 2055–2064. DOI: 10.1002/jmv.23905 | 2 | 2 |

**Table S6:**
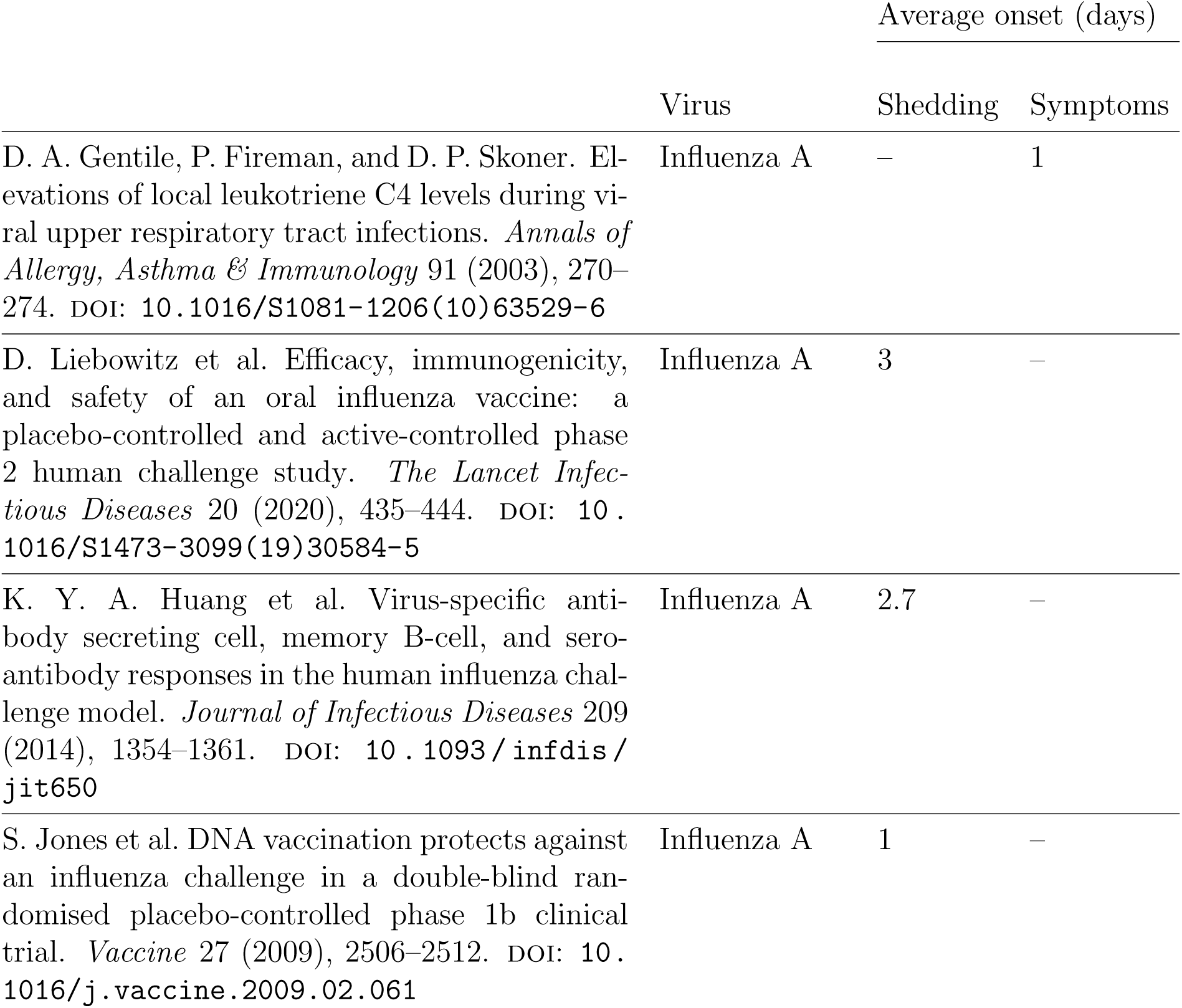
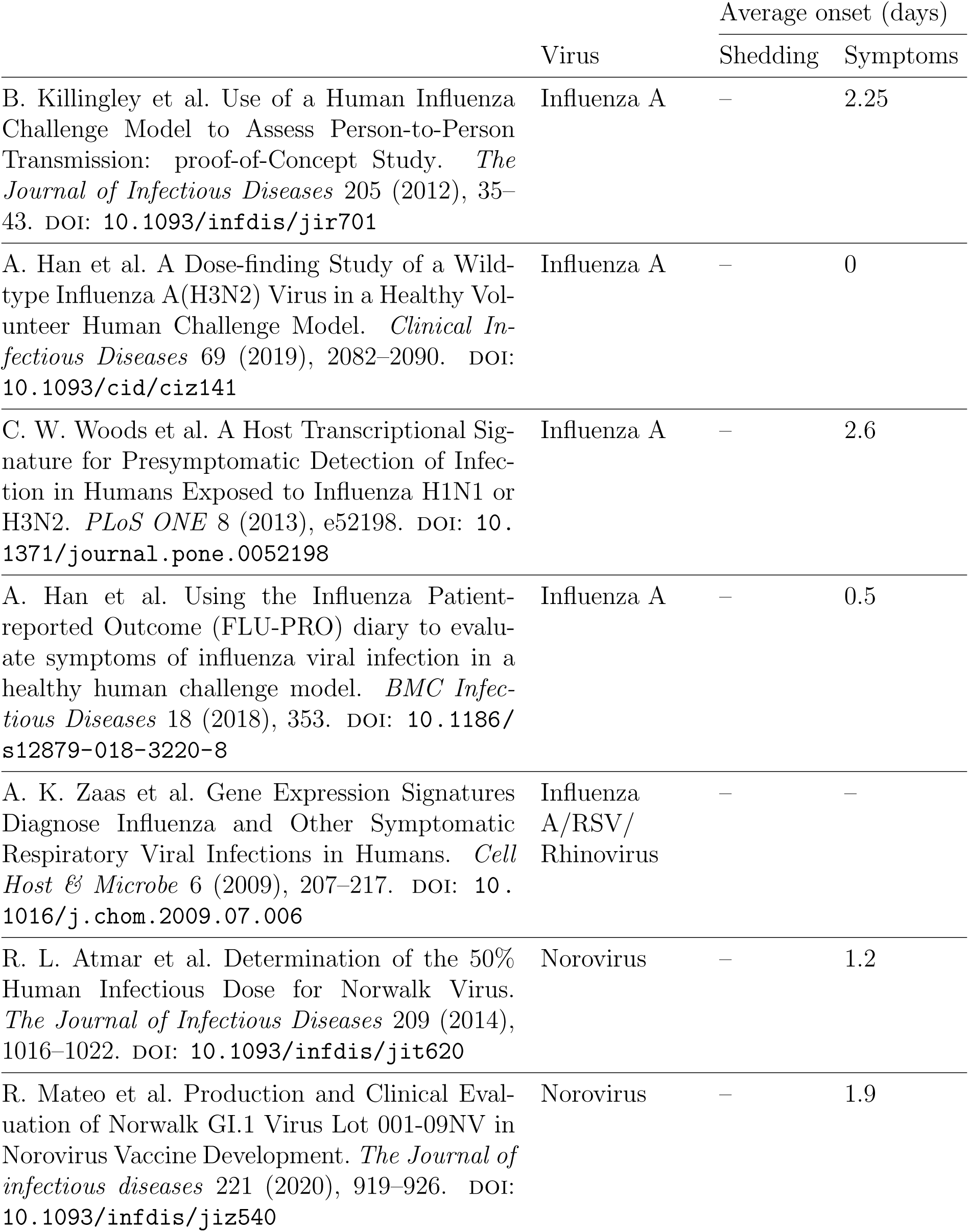
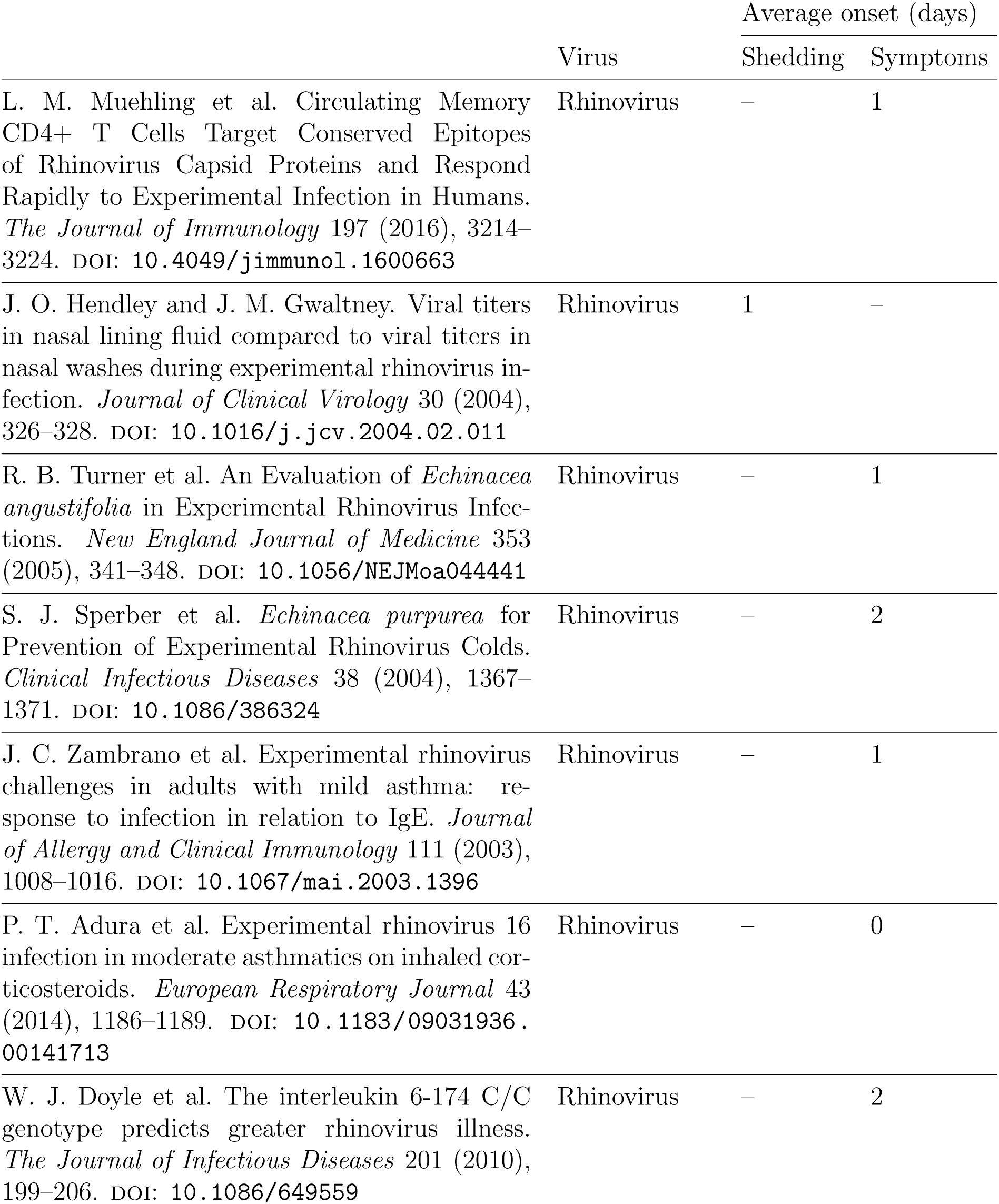
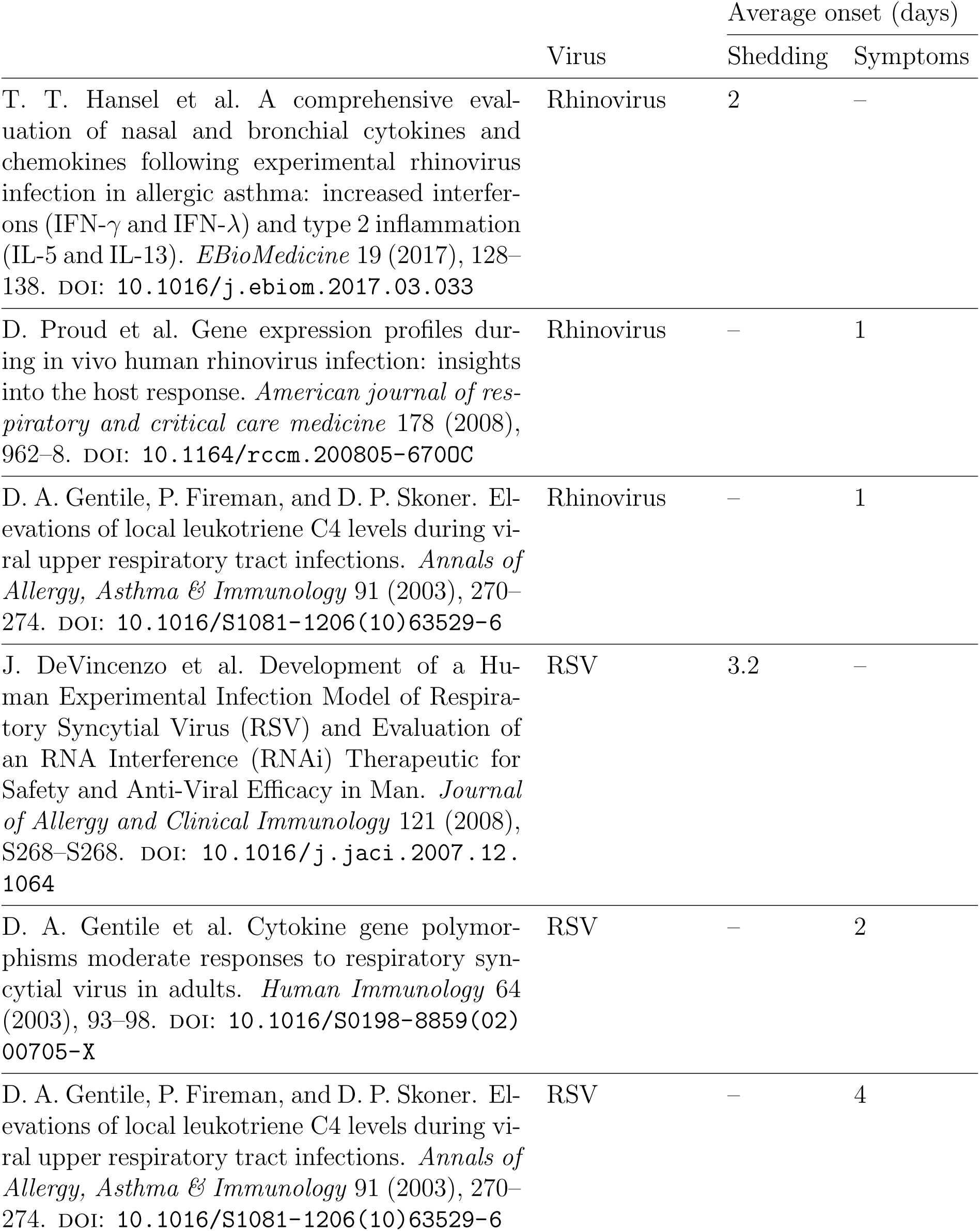
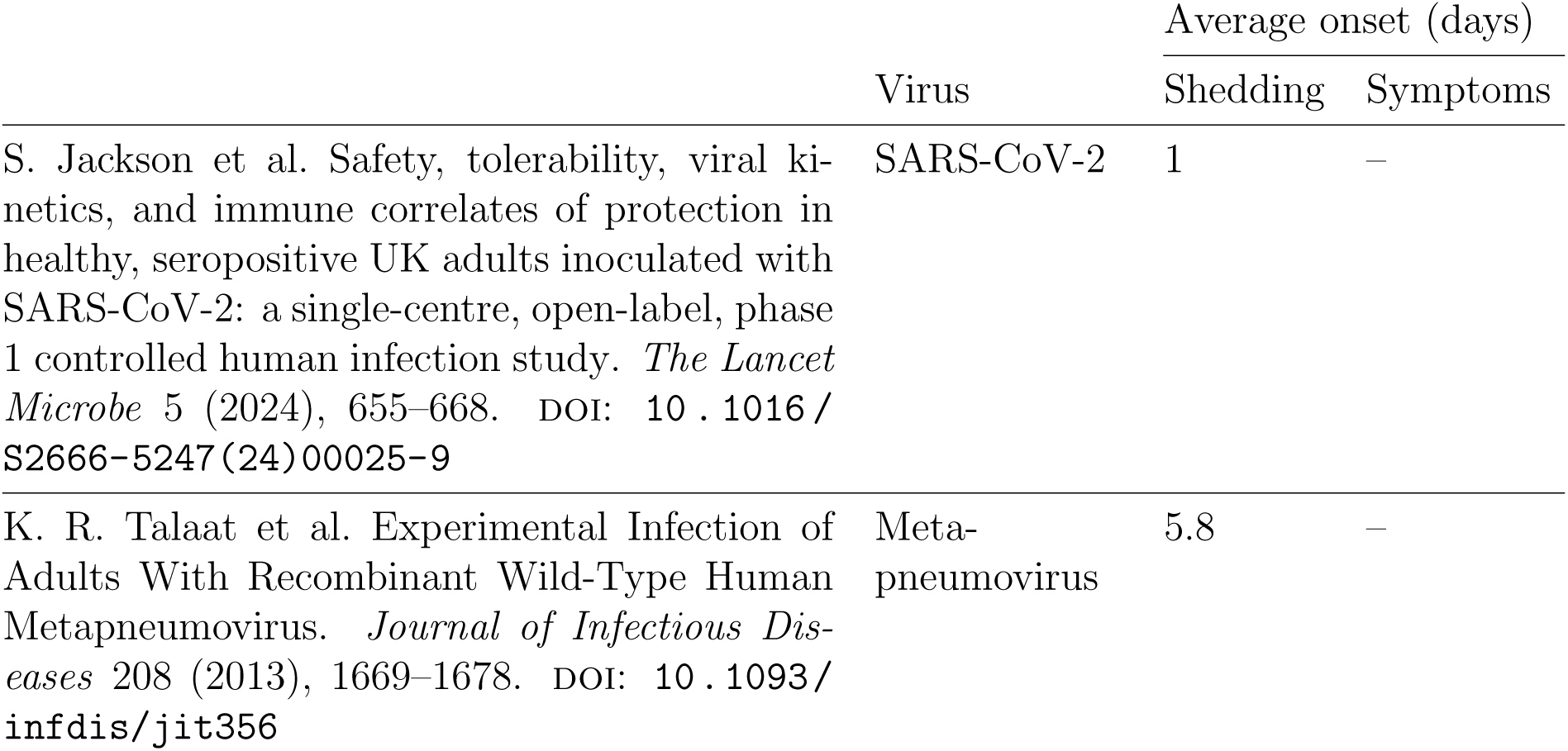
studies excluded from Figure 1. (‘–’ denotes unreported values)

**Table S7:**
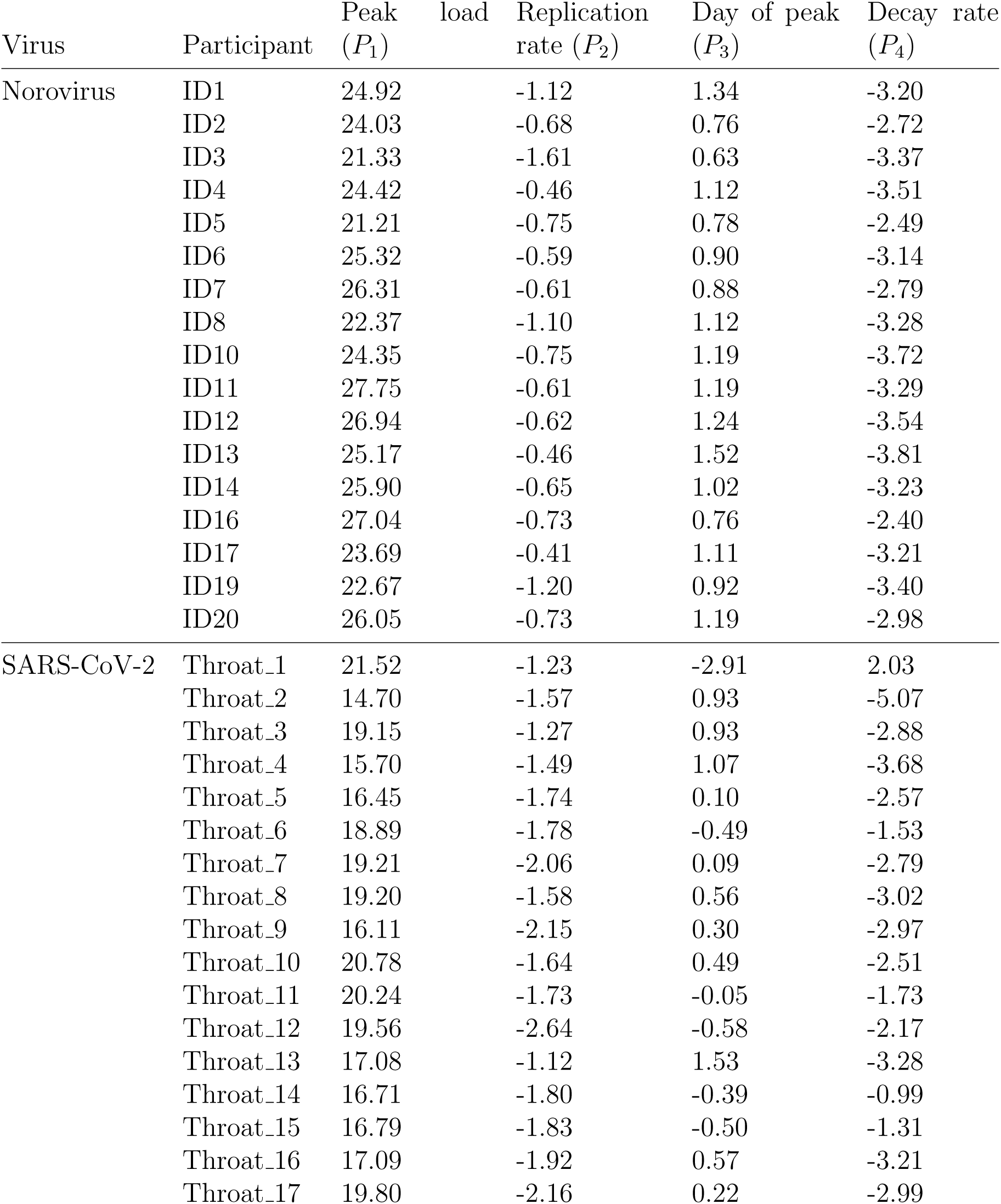
Fitted parameters for Equation 1 for each individual time series. Values are in the natural log scale.

**Table S8:**
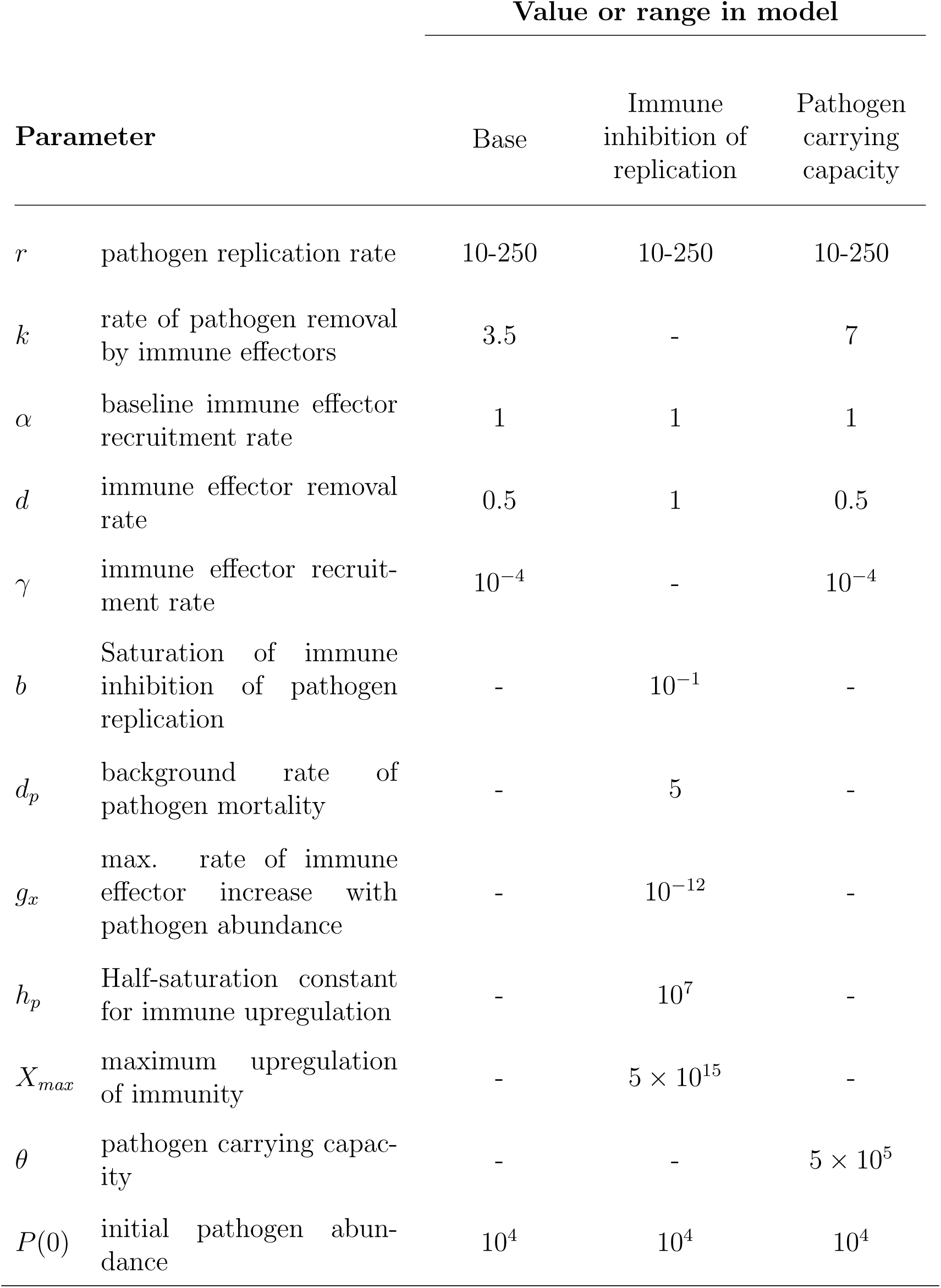
Parameters and values used for models (Eqns. 2-4).

|  |  | Value or range in model |  |  |
| --- | --- | --- | --- | --- |
| Parameter |  | Base | Immune inhibition of replication | Pathogen carrying capacity |
| $r$ | pathogen replication rate | 10-250 | 10-250 | 10-250 |
| $k$ | rate of pathogen removal by immune effectors | 3.5 | - | 7 |
| $\alpha$ | baseline immune effector recruitment rate | 1 | 1 | 1 |
| $d$ | immune effector removal rate | 0.5 | 1 | 0.5 |
| $\gamma$ | immune effector recruitment rate | $10^{-4}$ | - | $10^{-4}$ |
| $b$ | Saturation of immune inhibition of pathogen replication | - | $10^{-1}$ | - |
| $d_p$ | background rate of pathogen mortality | - | 5 | - |
| $g_x$ | max. rate of immune effector increase with pathogen abundance | - | $10^{-12}$ | - |
| $h_p$ | Half-saturation constant for immune upregulation | - | $10^7$ | - |
| $X_{max}$ | maximum upregulation of immunity | - | $5 \times 10^{15}$ | - |
| $\theta$ | pathogen carrying capacity | - | - | $5 \times 10^5$ |
| $P(0)$ | initial pathogen abundance | $10^4$ | $10^4$ | $10^4$ |

## Notes

### Competing Interest Statement

The authors have declared no competing interest.

### Funding Statement

This work was supported by the Cornell University College of Agricultural Sciences and the Cornell Institute of Host-Microbe Interactions & Disease Undergraduate Research Experience.

### Author Declarations

SARS-CoV-2 data were requested and obtained from the authors of Zhou et al. 2023 Lancet Microbe, a study approved by the UK Health Research Authority Ad Hoc Specialist Ethics Committee (reference: 20/UK/0002). All norovirus data were publicly available, save for total sample weights requested and obtained from Dr. Andreas Handel, from Ge et al. 2023 Emerging Infectious Diseases (for which the clinical protocol was reviewed and approved by the institutional review boards of the Baylor College of Medicine and The Houston Methodist Hospital).

### Summary of Updates

We expanded our literature search to include more controlled human infection (CHI) studies, revealing substantial variation in the relative timing of symptoms and shedding within trials for a given viral agent, along with considerable individual variation in the few studies that report those data. We extended our analysis on individual-level data on shedding and symptom timing to include recent CHI trial data for SARS-CoV-2 (Zhou \textit{et al}. 2023 \textit{Lancet Microbe}), a useful comparison for our analysis of norovirus data, now using the high-resolution data reported by Ge \textit{et al}. 2023 \textit{EID}. These two highly divergent viruses exhibit the same key pattern we uncovered in our original study, that faster viral replication hastens peak shedding without altering the timing of symptom onset. While a tradeoff between the rate and duration of transmission during pre-symptomatic transmission underpins important evolutionary theory (e.g., Saad-Roy \textit{et al}. 2020 \textit{PNAS}), we find no evidence of such tradeoffs from SARS-CoV-2 or norovirus. We also expanded our treatment of plausible within-host models for pre-symptomatic transmission, further underscoring the need to consider resource limitation. Our revised study demonstrates how models add value to individual-level CHI data, uncovering---or ruling out---within-host dynamics that contribute to the rapid and silent spread of deadly infections.

